# Targeting community-level drivers of antimicrobial resistance in sub-Saharan Africa: the effect of a community-based intervention bundle on household transmission of Extended Spectrum Beta-lactamase-producing *E. coli* in rural Burkina Faso - a cluster randomised trial

**DOI:** 10.64898/2025.12.15.25342269

**Authors:** R Aizouk, Y Sibidou, D Valia, B Ingelbeen, L Campbell, Juste S Kouanda, A Welgo, PM Diagne, B Riems, L Hardy, E Wouters, M Meudec, MAB van der Sande, T Halidou, BS Cooper, E van Kleef, CABU-EICO study team

## Abstract

**Background:** In sub-Saharan Africa (sSA), invasive antimicrobial-resistant infections often originate from community-level acquisition. We assessed whether a behavioural intervention bundle targeting sub-optimal antibiotic use and hygiene practices reduced household-level acquisition of extended-spectrum beta-lactamase–producing *E. coli* (ESBL-E).

**Methods:** We conducted a cluster-randomised controlled trial in 22 village clusters in Nanoro district, Burkina Faso. We enrolled 12 randomly selected households per cluster to assess intervention impact on ESBL-E household-transmission. The intervention comprised three rounds at three-month intervals and combined WHO AWaRe–based educational feedback for formal and informal medicine providers with a community-wide WASH and antibiotic-use behaviour change campaign. Consenting household members provided stool samples before, during, and after intervention rollout, alongside a pre–post household WASH survey. We estimated intervention effects on ESBL-E acquisition using Bayesian Markov models. Cox frailty models assessed associations between WASH exposures and acquisition. ClinicalTrials.gov, NCT05378880.

**Findings:** Between Oct 11, 2022, and Feb 19, 2024, 1203 individuals were enrolled. At baseline, 57·3% (346/604) of control and 48·6% (291/599) of intervention household members were colonised. Pre-intervention acquisition incidence was 3·8 per 100 person-days (95% credible interval [CrI] 2·0–9·9) in the intervention group and 3·5 (95% CrI 1·8–9·6) in the control group. The intervention did not change the risk of ESBL-E acquisition in months 1–6 (hazard ratio [HR] 1·02, 95% CrI 0·78–1·31), while we estimated a reduction in ESBL-E acquisition from months 6–9 (HR 0·82, 95% CrI 0·56–1·14). Acquisition risk was higher in the rainy season (peak HR 1·73, 95% CI 1·49–2·00), while improved sanitation was associated with lower risk (HR 0·77, 95% CI 0·59–1·00).

**Interpretation:** Findings, though inconclusive, were consistent with a modest intervention-related reduction in ESBL-E incidence. Higher acquisition rates associated with the rainy season and poor sanitation highlight the need to tackle environmental drivers of AMR transmission in addition to antibiotic use in rural sSA.

**Funding:** JPI-AMR CABU EICO grant number: JPIAMR2021-053

**Research in context:** *Evidence before this study:* A systematic review of antimicrobial stewardship interventions in both community and hospital-settings published in 2019 found that only 23% (190/825) reported microbiological outcomes, underscoring a lack of evidence on how stewardship interventions affect antimicrobial resistance (AMR). Since then, this gap remains, and particularly for sub-Saharan Africa (sSA). Here, invasive AMR infections are frequently community-associated, with household transmission considered a dominant pathway for community-level AMR acquisition. Two systematic reviews on community-level transmission of AMR bacteria, including one published in 2025, reported ESBL-E acquisition rates of 0·17–0·29 per 100 person-days, with half of individuals clearing their colonisation within 3–4 months. However, all of the included 11 studies were from high-income contexts. The Drivers of Resistance in Uganda and Malawi (DRUM) study provided important One Health insights into human, animal, and environmental reservoirs of ESBL-E and Klebsiella pneumoniae, and found high ESBL-E prevalences up to 60%. This study however, was observational and did not evaluate interventions, and to date, has not quantified community-level transmission. The published community-level interventions in sSA have largely focused on formal healthcare providers or prescribers to reduce sub-optimal antibiotic use. A recent scoping review identified only seven intervention studies targeting general communities in sSA, highlighting a lack of rigorous evaluations of community-centred stewardship and AMR mitigation efforts more broadly and outside formal healthcare settings. None of these studies measured microbiological outcomes or effects on community transmission of resistant bacteria.

*Added value of this study:* The CABU-EICO trial is, to our knowledge, the first cluster-randomised AMR intervention in a rural, low-income setting to quantify the effects of an intervention targeting both providers and communities on community-level ESBL-E transmission-dynamics. Using repeated stool sampling from household members and a continuous-time multi-state modelling framework, we estimated household-level ESBL-E acquisition and duration of colonisation, and found evidence for a reduction in ESBL-E transmission following the intervention. Additionally, we quantified seasonal patterns in the risk of ESBL-E acquisition in our West African setting, showing a peak during the rainy season, despite reportedly lower antibiotic use during this time of year. By moving beyond prevalence-based outcomes and antibiotic-use metrics alone, our intervention evaluation provides a statistically more efficient and mechanistically informative framework for evaluating AMR interventions.

*Implications of all the available evidence:* This study shows that household-level transmission of ESBL-E is substantial in rural sSA and markedly higher than estimates from high-income settings, with clear seasonal peaks during the rainy season and increased risk associated with poor sanitation. Together with recent One Health genomic evidence demonstrating frequent transmission between humans, animals and the environment in Eastern Africa, these findings suggest that community-level AMR dynamics are driven by both antibiotic selection pressure and environmental exposure pathways. Effective AMR control in similar settings will therefore require, similar to our approach, integrated One Health strategies that combine antibiotic stewardship, with structural and environmental interventions, and that incorporate transmission as well as acquisition outcomes to fully capture intervention impact.

## Introduction

Antimicrobial resistance (AMR) is considered a critical global health threat, disproportionately affecting resource-limited settings, sub-Saharan Africa (sSA) in particular.^1,2^ Key assumed drivers contributing to the propagation of AMR include inappropriate use of antibiotics, poor Water, Sanitation, and Hygiene (WASH) conditions, and close contact with animals and/or food products, particularly in settings characterised by a low socioeconomic position.^3,4^ Recognising the urgency of this threat, world leaders committed in September 2024 to ambitious targets to curb AMR, including reducing AMR-related mortality by 10% by 2030, achieving 70% global use of Access antibiotics, and ensuring universal access to improved WASH standards.^5^

In sSA, invasive AMR syndromic infections are often caused by pathogens associated with community-level acquisition.^6^ Extended-spectrum-lactamase-producing *Escherichia coli* (ESBL-E) serves as a commonly used proxy for AMR spread, given its widespread occurrence and its frequent identification as causative agent of severe infections globally.^7^ Invasive infections caused by AMR bacteria, including ESBL-E, are linked to asymptomatic colonisation, with human-to-human transmission within households considered an important yet understudied pathway both in low- and high income settings.^8–10^,^11–12^ A recent study from urban Nairobi identified within-household transmission as a predominant source of ESBL-E strain sharing.^12^ Other studies from rural sSA, including Nanoro (Burkina Faso), and Malawi, have reported high colonisation rates with ESBL-E in healthy individuals, which varied by rainy and dry seasons, and were associated with household environmental exposures including clean water access, and hand hygiene practices.^13,14^ Misuse and overuse of antibiotics could increase intestinal carriage of AMR bacteria, hence raising colonisation pressure and facilitating onward transmission, especially in settings where hygiene standards are poor, and self-medication is common.^15,16^

Despite evidence pointing to household and community spread of AMR bacteria, interventions to mitigate AMR in resource-limited settings have predominantly focused on healthcare settings.^11^ Reported stewardship interventions, including those from LMICs, have shown reductions in antibiotic use across both inpatient and outpatient/community settings. However, these studies generally suffer from low methodological quality and report variable effects on improving antibiotic use, in part related to heterogenous intervention designs.^17,18^ Among randomised controlled trials of community-level stewardship interventions in LMICs, only one study from India targeted informal medicine sellers. Bundled stewardship strategies, which combine persuasive and/or educational approaches targeted at the community, generally showed most promise in improving antibiotic use.^18,19^ However, the impact of these community-based interventions on microbiological outcomes - such as reducing acquisition of AMR bacteria - as well as clinical outcomes are rarely measured or reported.^17^

To address these evidence gaps, we conducted a cluster-randomised controlled behavioural intervention trial in rural sSA to evaluate the effect of an intervention bundle, co-developed with healthcare providers and community members from rural Burkina Faso and Democratic Republic of Congo (DRCongo). The intervention targeted community-level medicine providers, and surrounding communities to reduce sub-optimal antibiotic use and improve hygiene.^20^ We evaluated both the effect of the intervention on community-level antibiotic use - reported in^21^] - and its impact on within-household transmission of bacterial AMR, reported here.

## Methods

### Study design

CABU-EICO study was implemented in both Burkina Faso and DRCongo as a multi-country cluster-randomised controlled trial. The present paper reports on the microbiological results from the Burkina Faso study site where we conducted longitudinal stool sampling to evaluate the intervention effect on AMR acquisition and decolonisation dynamics. Located in the Nanoro Health District, the latter site has established field and laboratory infrastructure, prior evidence from pilot work of high prevalence of community ESBL-E carriage ^22^,and, thanks to a Health and Demographic Surveillance System (HDSS), well-characterised household structures that allowed longitudinal follow-up. Eligible clusters consisted of villages with ≥500 residents with at least one medicine outlet, i.e. primary health centre or informal community-level medicine provider as the main medical dispenser. The study protocol was approved by the Ethics Committee for Health Research of Burkina Faso (reference N°2022-03-050) and obtained from the Institutional Review Board of the Institute of Tropical Medicine, Antwerp, Belgium (1559/22, dd 29/03/2022), and from the Ethics Committee of the Antwerp University Hospital, Belgium (3363, dd 09052022).

### Randomisation and masking

In total, 22 clusters were randomised 1:1 to either intervention or control arms (Figure S1). We stratified randomisation by village cluster community-level medicine provider (i.e. government run health centre vs other formal or informal medicine provider). Total population coverage in the 22 clusters was 82,018 individuals. To evaluate ESBL-E acquisition/decolonisation and WASH outcomes related to the intervention, we randomly selected 36 households per cluster from the HDSS, and conducted stool sampling in a subsequent twelve randomly selected households.

### Intervention and participants

The intervention is described in detail in Table S1. Briefly, the intervention bundle was implemented, for each cluster, over a six-month period, covering three rounds three months apart. Each of the 11 intervention village clusters were subsequently visited for 2-4 days per round, and followed up for nine-months (Figure S1). A number of enabling and persuasive interventions were co-developed following qualitative focus groups, photovoice and interviews with local community members and formal and informal medicine providers over a six-month period. These interventions targeted suspected main modifiable drivers of AMR, i.e. unnecessary antibiotic use and WASH (Table S1), combining provider-focused antimicrobial stewardship education based on the WHO AWaRe Antibiotic Book with community-wide WASH, and antibiotic-use behaviour change education activities (Table S1). The first two rounds introduced adapted treatment guidance for four syndromes accounting for most community antibiotic use, while the third round reinforced prior content and focused on provider–patient communication on antibiotic use and community commitments to appropriate care-seeking and hygiene practices.

To evaluate ESBL-E acquisition/decolonisation and WASH outcomes related to the intervention we included households and household members using a household stool collection survey and a WASH survey (supplementary material section 1) among those meeting the following inclusion criteria (box 1).

#### Household stool collection

Stool collection took place between 11 October 2022 and 19 February 2024. Stool samples were collected from all eligible and consenting household members three months before the start of the intervention, at the start, and at month 3, and 9 post-intervention start in both intervention and control clusters using pre-labelled sterile containers with an electronic questionnaire (Figure S1, supplementary material section 1). Written informed consent was obtained from all individuals aged at least 18 years. For patients aged between 14 and less than 18 years, oral assent was obtained in addition to parents or caretakers’ written informed consent. For patients under 14 years, written informed consent was obtained from parents or caretaker. Faecal sampling was undertaken from all consenting household members. In case of more than six household members, faecal samples were collected from six randomly selected individuals. Field workers provided instructions to household members on sampling and storage. The following morning, stool samples were collected by trained field agents, and the specimens were transported to the laboratory in a cooler box maintaining temperatures between 2–8 °C. All samples reached the clinical microbiological laboratory in Nanoro within 8 hours of production, in accordance with procedures ensuring biosafety for field staff and the public. Upon arrival at the laboratory, each sample was recorded and processed immediately on selective CHROMagar™ ESBL plates according to the manufacturer’s instructions. Suspected ESBL-producing *E. coli* were further identified using standard biochemical tests. Antimicrobial susceptibility testing was performed following CLSI guidelines.

#### Household WASH survey

A household WASH survey took place from 11 October 2022 - 1 February 2023 (baseline), and 12-months later (post-intervention round). At baseline, household member heads or another available adult household member were surveyed using an electronic questionnaire recording details on household structure, WASH exposures following UNICEF and WHO guidance,^22^ in addition to antibiotic consumption in the last month and last three months and healthcare-seeking behaviours for all household members.

### Outcomes

The first primary outcome of this trial, changes in provision of WHO Watch-group antibiotics is reported in the joint submission. Here we report in detail on the second primary outcome, i.e. change in rates of person-to-person transmission and duration of carriage of ESBL-producing *E. coli* within households. We assessed these outcomes as:

1. Change in instantaneous acquisition and decolonisation hazards, expressed as the intervention hazard ratio for ESBL-E acquisition/decolonisation, capturing how the intervention altered the rate at which susceptible individuals acquired/cleared ESBL-E colonization.
2. Change in acquisition incidence density, expressed as the relative reduction in ESBL-E incidence per 100 person-days. This captures how the intervention altered cumulative population-level ESBL-E acquisition over the full follow-up period using the incidence rate ratio (IRR).

For both measures, we summarised posterior distributions by reporting the probability of a reduction (HR or IRR < 1), the probability of an increase (HR or IRR > 1), and the probability of a meaningful reduction (HR or IRR < 0.9). Further secondary outcomes included the change in hygiene practices and exposures (reported here for the Nanoro site only).

### Statistical analyses

#### Sample size calculation

For the stool sample collection, using a simulation-based approach, we estimated the power to detect reductions in transmission resulting from the intervention (supplementary material section 2). For an intervention that reduces the transmission rate by 40%, 30%, and 20% respectively, and with a two-sided type 1 error of 5% we estimated that a study with 12 households per cluster and 22 clusters would have 99%, 82% and 43% power respectively to detect a reduction in transmission. This implied 132 households per intervention arm. For a change in WASH indicators, sample size calculation assumed that 30% of households practiced correct handwashing at baseline. With an intra-cluster correlation coefficient of 0.05 and a design effect of 2.75, a sample of 792 households across 22 clusters (36 per cluster) provided 80% power to detect a 15-percentage-point absolute increase in correct handwashing (e.g., from 30% to 45%) with 80% power and an expected precision of ±5.3 percentage points for baseline estimates.

#### Intervention evaluation

We evaluated multiple intervention-effect scenarios to reflect plausible mechanisms through which the intervention could influence ESBL-E acquisition dynamics. In the baseline scenario (scenario 1), we assumed a progressive intervention effect, whereby the first two rounds - introducing provider-focused antimicrobial stewardship and community behaviour change activities - reduced acquisition rates during an early intervention phase (months 1–6), with the third round reinforcing prior messages and potentially yielding additional reductions during a late intervention phase (months 6–9).

In alternative intervention effect scenarios we:

- Assumed a single immediate sustained intervention effect across the study period, consistent with the later rounds maintaining rather than enhancing intervention effects (scenario 2),
- Extended scenarios 1 and 2 by assuming that the intervention could affect both acquisition and decolonisation hazards, i.e. shortening colonisation duration in addition to reducing acquisition risk (scenario 3 and 4).

Measured change in ESBL-E acquisition dynamics.

To evaluate the effect of the intervention bundle on ESBL-E acquisition and decolonisation, we fitted Bayesian continuous-time Markov chain (CTMC) models to the longitudinal stool data. This framework jointly estimates the individual-level acquisition rate (λ□□), the loss-of-carriage rate (λ□□), and how these rates vary with the intervention and covariates at individual and household levels. Each participant is assumed uncolonised (S_1_) or colonised with detectable ESBL-E (S_2_), and transitions between these states are determined by the underlying hazard rates (Figure S1B). The model incorporated seasonality, demographic factors (age, sex), and household-level clustering, with normal distributed random effects capturing between-household variation in not explicitly included AMR drivers. Posterior summaries of the between-household variance were obtained from the posterior distribution. Unlike standard time-to-event models, the CTMC approach infers transitions by integrating hazards over each observation interval, allowing for uncertain event times and multiple transitions that may occur between sampling points. Because intervention rounds were implemented at different times across villages, the model accounted for village-specific intervention start dates, enabling correct attribution of exposure within each household’s observation intervals for the early intervention effect (after the start of intervention rounds 1 and 2) and late intervention effects (after intervention round 3). To capture temporal fluctuations in acquisition risk, we estimated daily transition rates across 28-day calendar-time segments. Seasonal structure was represented using both a sinusoidal term (Scenarios 1-4A) and a penalised B-spline smooth over calendar time (Scenarios 1-4B). We assessed comparative model fit of models with and without terms for seasonality and different intervention impact scenarios (Table S2) using leave-one-out cross-validation (LOO-CV). All models were implemented in *Stan version 2.23.2*. Further modelling detail, including model fitting, rigorous simulation-based model checking procedures, as well as sensitivity analyses are provided in Supplementary Material Section 2.

#### WASH exposures associated with ESBL-E acquisition risk

As a contextual analysis to understand which intervention-related changes in AMR drivers might explain the estimated effect on ESBL-E acquisition, we first assessed whether WASH-related exposures were associated with the risk of ESBL-E acquisition in our study site. We fitted mixed-effects Cox proportional hazards frailty models for each predefined WASH indicator, estimated using a staggered-entry (left-truncation) framework on a calendar-time scale (days since the global study start date). Because the intervention also targeted antibiotic use - an important potential mediator of ESBL-E acquisition that was not the focus of this analysis and for which detailed household- or cluster-level data were not available - we restricted this analysis to participants in the control arm. For individuals who converted from ESBL-E negative to ESBL-E positive between sampling visits, the event time was imputed as the midpoint between the last negative and first positive sample. Those who remained ESBL-E negative were censored, as well as those who went from positive to negative were censored at their last observation. Each participant contributed person-time from the date of their first ESBL-E negative sample. To account for clustering, random intercepts were included at both the individual and household levels. The model was implemented in R (version 4.4.1) using the *coxme* package.

#### Measured change in WASH exposures

We developed a binary WASH evaluation framework in collaboration with a WASH expert (BR), drawing on WHO/UNICEF Joint Monitoring Programme definitions,^23^ and validated indicator coding with DV to ensure contextual appropriateness. This allowed for estimating population-weighted prevalence ratios (PRs) for six WASH indicators targeted by the intervention community-campaign, capturing access to improved drinking water, sanitation, correct handwashing, and livestock-related exposure risks (Table 1; coding in Table S2). Using household survey data, we estimated intervention effects with survey-weighted quasi-Poisson regression models with a log link and an interaction between intervention group and survey round. Models accounted for household clustering within villages, sampling weights proportional to village population size divided by the number of households surveyed per round, and rainy season to capture seasonal variation in WASH behaviours. To allow comparison across villages surveyed at different times, we standardised predicted prevalences to the dry season. We conducted all analyses in R (version 4.4.1) using the *survey* package.

**Table 1:** Baseline characteristics of study population from households where follow-up faecal samples were collected.

|  |  | **Control**<br>**(N=604)** | **Intervention**<br>**(N=599)** | **Total**<br>**(N=1203)** |
| --- | --- | --- | --- | --- |
| **Household characteristics** |  |  |  |  |
| N households |  | 133 | 132 | 265 |
| N households with individuals with >1 observation (%) |  | 131 (98·5%) | 128 (97·0%) | 259 (97·7%) |
| N household per cluster, median [min, max] |  | 12 [12 - 12] | 12 [12 -13] | 12 [12 - 13] |
| Household size, median [min, max] |  | 8·00 [3·00, 37·0] | 8·00 [1·00, 32·0] | 8·00 [1·00, 37·0] |
| N household members tested per household, median [IQR] |  | 4 [3 - 6] | 4 [3 - 6] | 4 [3 - 6] |
| % Children <5, median [min, max] |  | 1·00 [0, 9·00] | 2·00 [0, 10·0] | 2·00 [0, 10·0] |
| **Individual characteristics** |  |  |  |  |
| N individuals with >1 observation (%) |  | 582 (96·4%) | 569 (95·0%) | 1097 (91·2%) |
| N individuals with 4 observations (%) |  | 390 (64·6%) | 357 (59·6%) | 747 (62·1%) |
| Age, median [min, max] |  | 18·0 [1·00, 92·0] | 18·0 [1·00, 85·0] | 18·0 [1·00, 92·0] |
| Age group, n (%) |  |  |  |  |
|  | 0-4 years | 86 (14·2%) | 73 (12·2%) | 159 (13·2%) |
|  | 5-17 years | 215 (35·6%) | 243 (40·6%) | 458 (38·1%) |
|  | 18-64 years | 185 (30·6%) | 188 (31·4%) | 373 (31·0%) |
|  | 50+ years | 113 (18·7%) | 84 (14·0%) | 197 (16·4%) |
|  | Missing | 5 (0·8%) | 11 (1·8%) | 16 (1·3%) |
| Sex, n (%) |  |  |  |  |
|  | Male | 241 (39·9%) | 244 (40·7%) | 485 (40·3%) |
|  | Female | 363 (60·1%) | 352 (58·8%) | 715 (59·4%) |
|  | Missing | 0 (0%) | 3 (0·5%) | 3 (0·2%) |
| Social economic status |  |  |  |  |
|  | Highest quintile | 111 (18·4%) | 72 (12·0%) | 183 (15·2%) |
|  | Fourth | 79 (13·1%) | 119 (19·9%) | 198 (16·5%) |
|  | Third | 148 (24·5%) | 102 (17·0%) | 250 (20·8%) |
|  | Second | 126 (20·9%) | 222 (37·1%) | 348 (28·9%) |
|  | Lowest | 128 (21·2%) | 46 (7·7%) | 174 (14·5%) |
|  | Missing | 12 (2·0%) | 38 (6·3%) | 50 (4·2%) |

#### Handling of missing data

In all analyses, for age and sex, missing entries were imputed using the follow-up observation closest in time. Individuals with no age or sex recorded at any time were excluded. Sampling dates were taken from stool collection records, with consent dates used as proxies when missing. For individuals with no ESBL-E stool collection at baseline (n=17), we imputed the ESBL-E status by sampling from a Bernoulli distribution using the baseline prevalence specific to their trial arm as the probability. The CTMC was fitted to data from individuals with at least two observations; participants with fewer than two valid observations were excluded because transitions could not be modelled. The Cox proportional hazard model was fitted to individuals with four stool samples, to allow for equally spaced observation intervals. All other individuals were excluded.

## Results

### Description of the study population

Of the 1203 individuals enrolled at baseline, 1097 (91·2%) from 259/265 households provided at least two consecutive stool samples. Households were generally large, with a median size of 8 members [IQR: 6–12]. A median of four household members were tested per household [IQR: 3–6].

The study population was predominantly female (60·1%, 723/1203), with a median age of 18 years [range: 1–92 years] and a majority (373/1203, 31·0%) between 5-17 years old (Table 1). WASH characteristics, measured in 808 households, were comparable for handwashing, and animal faeces identified on household floors (Table S3).

Differences were only observed in primary drinking water sources. In the control arm, 336/409, 82·2% (dry) and 324/409, 79·2% (rainy) of households used an improved source, compared with 383/399, 96·0% and 367/399, 92·0% in the intervention arm. Similarly, sanitation practices differed between intervention groups, with unimproved sanitation reported in 266/409, 65·0% control, versus 313/399, 78·4% intervention households. These unimproved sanitation practices largely concerned open defecation, while improved primary water sources were mostly boreholes. Animal contact was widespread, occurring both inside and outside household structures (382/409, 93·4% and 349/399, 87·5% of the control and intervention households respectively).

### Pre-intervention ESBL-E transmission dynamics

At baseline, 346/604 (57·3%) of individuals were identified positive for ESBL-E colonisation in the control group. This was 291/599 (48·6%) in the intervention group (Table S4 shows prevalence rates by randomisation strata, revealing no source of imbalance). Under the base case scenario, our model estimated a baseline daily acquisition rates of 0·0135 (95% CrI: 0·0095 - 0·0239) and an ESBL-E loss-of-carriage rates of 0·0095 (95% CrI: 0·0065–0·0147), corresponding to a median duration of colonisation of ∼3·4 months (102 days [95% CrI: 68–154]). Household-level variance in the log-hazard scale was estimated at 1·82 (95% CrI: 1·13–2·78), corresponding to a standard deviation of 1·35 (95% CrI: 1·07–1·67), indicating substantial heterogeneity in transition rates across households (Table S5). Pre-intervention ESBL-E incidence rates were comparable between control and intervention villages, at 3·8 (95% CrI: 2·0–9·9) and 3·5 (95% CrI: 1·8–9·6) per 100 person-days, respectively. Incidence rates among villages varied from 2·6 (95% CrI: 1·0–17·8) to 4·7 (95% CrI: 1·9–17·1) per 100 person-days, with one village estimated to have a markedly lower baseline rate (95% CrI: 0·1 [0·1–0·9], Table S6).

### Intervention bundle effect on ESBL-E transmission dynamics - Base case scenario

Over the 9-month period after the intervention-start, ESBL-E acquisition incidence averaged 7·4 per 100 person-days in control villages (95% CrI: 3·9–18·2) and 6·3 per 100 person-days in intervention villages (95% CrI: 3·2–17·2), yielding an incidence rate ratio (IRR) of 0·91 (95% CrI: 0·67–1·30).

We observed no reduction in the acquisition hazard during the early intervention phase (months 1–6; hazard ratio [HR] 1·02, 95% CrI: 0·78–1·31), with posterior probabilities close to equipoise (i.e. 45% probability for a reduction; 18% for a meaningful reduction). In the later intervention phase (months 6–9), there was some evidence of a decline in acquisition hazard in the intervention villages (HR 0·82, 95% CrI: 0·56–1·14), corresponding to an 88% probability of a reduction and a 71% probability of a meaningful reduction (HR < 0.9, Figure 3, Table S7). Intervention effects were broadly consistent across the 11 intervention villages, with late-intervention phase ESBL-E IRRs relative to controls ranging from 0·71 (95% CrI: 0·37–1·15) to 0·81 (95% CrI: 0·50–1·32, Table S6).

**Figure 1:**
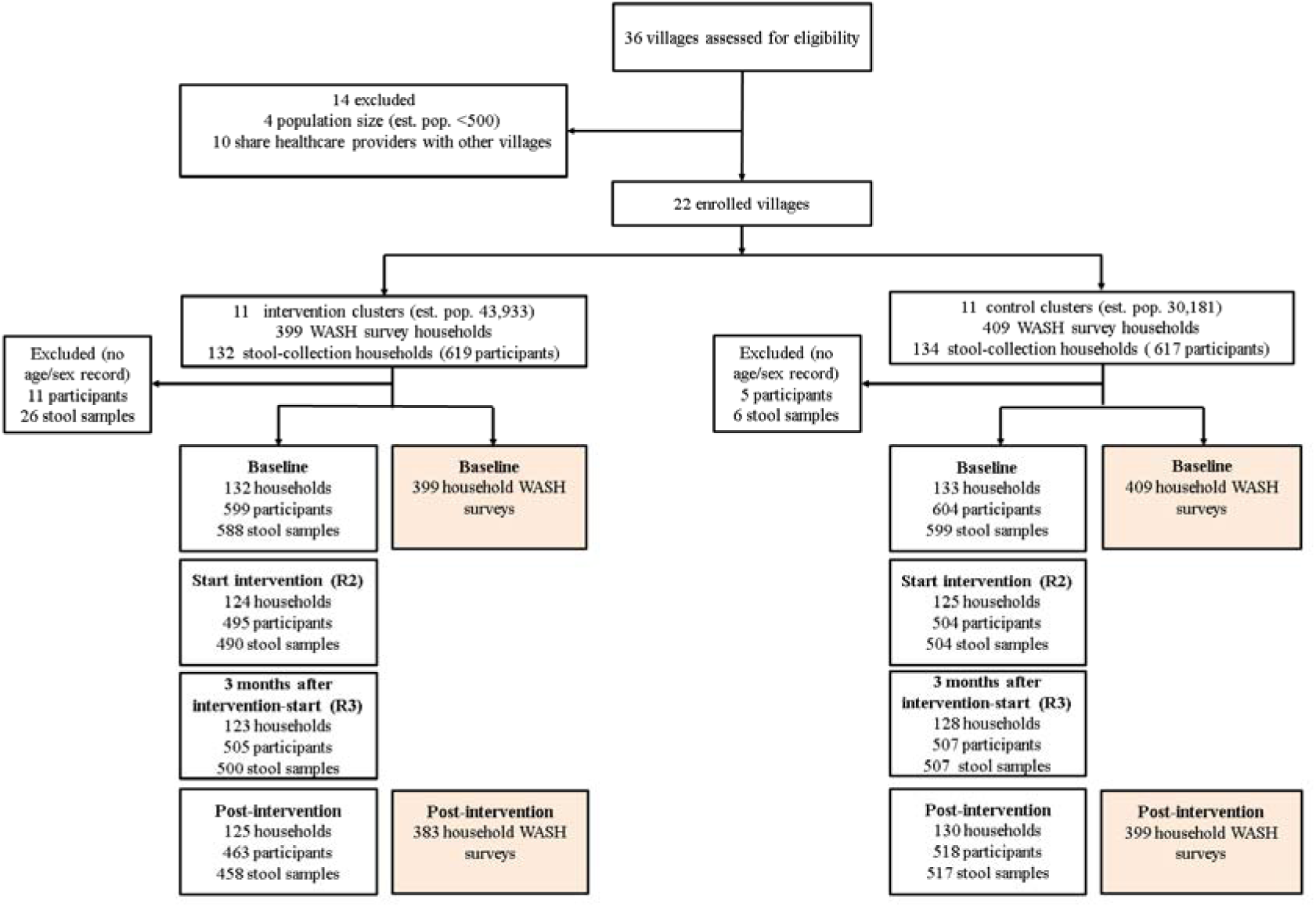
Trial profile diagram. Villages were assessed for eligibility, enrolled, and allocated to intervention or control clusters. The diagram shows the number of households, participants, and stool samples contributing data across study rounds, including baseline, intervention start, three months after intervention start, and post-intervention. Exclusions due to village eligibility criteria and missing age or sex information are indicated for each intervention group. WASH = water, sanitation, and hygiene. R2 = intervention start; R3= three months after intervention start.

**Figure 2:**
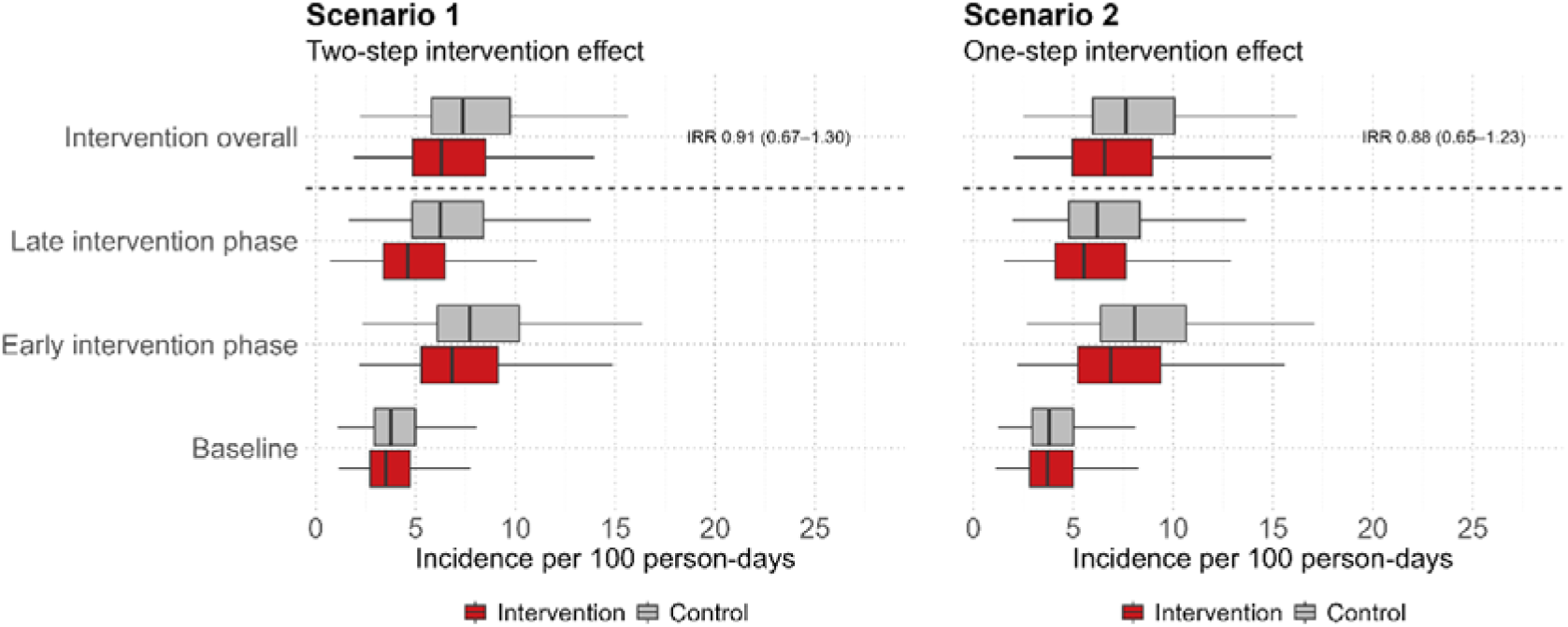
Intervention effect on ESBL-E acquisitions. ESBL-E acquisition is expressed as incidence per 100 person-days per intervention phase of the intervention vs control group for scenario 1 (A) and scenario 2 (B). In scenario 1, a separate effect for the early post-intervention phase (1-6 month post-intervention) and the late post-intervention phase (6-9 months) was estimated. In scenario 2, a constant intervention effect was estimated, assuming the different intervention rounds would maintain the effect.

**Figure 3:**
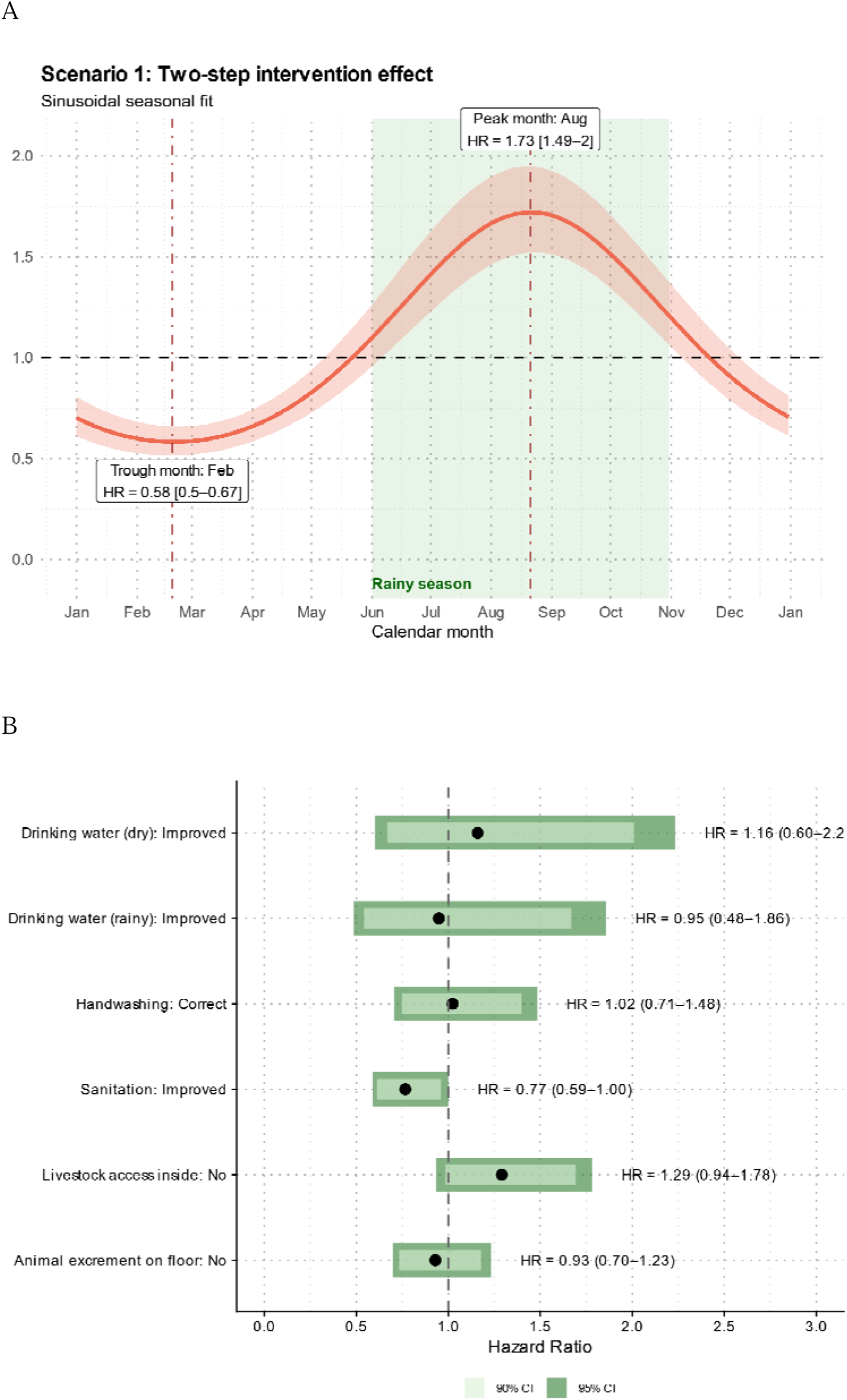
Seasonal pattern of ESBL-E acquisition (A). Baseline, sinusoidal approach. **Effect of individual intervention targets on acquisition from the Cox proportional hazard models (B)**. Hazard ratio for the risk of household acquisition of ESBL-E in rural Burkina Faso among individuals in the control group. Shadings represent from light to dark: 95% Confidence Intervals (CI), 80%CI and 50%CI respectively. HR<1 indicates a decreased risk, HR>1 indicates an increased risk.

### Intervention bundle effect on ESBL-E transmission dynamics - model scenario analyses

Base case intervention scenario 1 had a comparable model fit to intervention scenario 2 (Table S8), with good predictive performance (Figure S2), and similar estimate for between-household variance (Table S5). Under the latter scenario, assuming a consistent intervention effect across the 9-month intervention period, the intervention was similarly associated with a modest reduction in ESBL-E acquisition incidence (IRR 0·88 (95% CrI: 0·65 - 1·23, Figure 3). In contrast, in scenarios that allowed the intervention to affect both acquisition and decolonisation hazards (scenarios 3 and 4), there was strong correlation between acquisition and decolonisation parameters in the posterior distribution (Figure S3) and poor convergence (Table S8). These patterns indicate identifiability limitations, suggesting that the available data cannot reliably separate intervention effects on acquisition from those on decolonisation.

### Seasonal variation in ESBL-E transmission dynamics

ESBL-E incidence rates increased in both intervention and control groups over the study period (Figure 3A), which was explained by a pronounced seasonal pattern, with increased ESBL-E acquisition during the rainy season, with a peak acquisition risk in August (HR 1·73, 95% CrI: 1·49–2·00) - and lowest risk in February (HR 0·58, 95% CrI: 0·50–0·67).

Fitting a more flexible cubic B-spline showed a similar seasonal peak and trough timing. Of note, the latter model exhibited less stable convergence and some divergent transitions (Table S4).

### WASH practices associated with ESBL-E acquisition

Among control arm participants, 390/604, 63.2% provided four stool samples, with a baseline ESBL-E colonisation prevalence comparable to that of the full control study population (57·3%). Improved sanitation was associated with a lower risk of ESBL-E acquisition (HR 0·77, 95% CI 0·59–1·00; Figure 3B). No other WASH exposures and behaviours showed strong associations with acquisition risk.

### Intervention related changes in WASH practices

The intervention did not convincingly improve household-level WASH behaviour (Table 2). Over the 12-month period, hygiene indicators related to modifiable behaviour targeted in the intervention showed little to no change, and trends were similar in both arms.

**Table 2:**
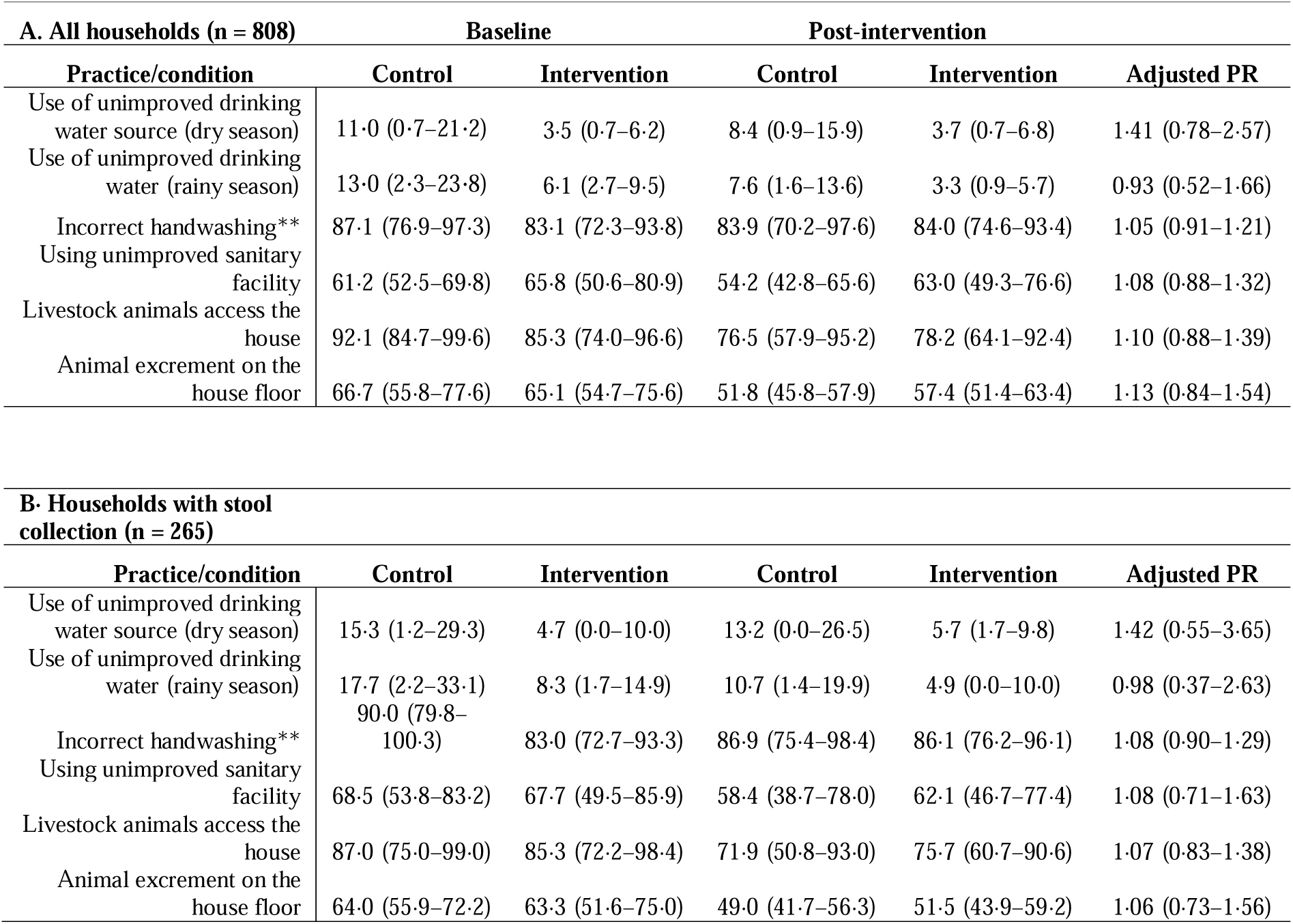
Change in WASH-indicators in all households where a household survey was taken (A) and households where a household survey and stool was sampled (B). Correct handwashing is defined as observed handwashing following defecation with soap available and used. Prevalence ratios (PRs) were estimated using survey-weighted quasi-Poisson regression models with a log link, accounting for household clustering within villages and survey weights proportional to the village population size divided by the number of households surveyed. Models included an interaction between intervention arm and survey round to estimate difference-in-differences effects.

| A. All households (n = 808) |  |  |  |  |  |
| --- | --- | --- | --- | --- | --- |
| Practice/condition | Baseline |  | Post-intervention |  | Adjusted PR |
|  | Control | Intervention | Control | Intervention |  |
| Use of unimproved drinking water source (dry season) | 11.0 (0.7–21.2) | 3.5 (0.7–6.2) | 8.4 (0.9–15.9) | 3.7 (0.7–6.8) | 1.41 (0.78–2.57) |
| Use of unimproved drinking water (rainy season) | 13.0 (2.3–23.8) | 6.1 (2.7–9.5) | 7.6 (1.6–13.6) | 3.3 (0.9–5.7) | 0.93 (0.52–1.66) |
| Incorrect handwashing** | 87.1 (76.9–97.3) | 83.1 (72.3–93.8) | 83.9 (70.2–97.6) | 84.0 (74.6–93.4) | 1.05 (0.91–1.21) |
| Using unimproved sanitary facility | 61.2 (52.5–69.8) | 65.8 (50.6–80.9) | 54.2 (42.8–65.6) | 63.0 (49.3–76.6) | 1.08 (0.88–1.32) |
| Livestock animals access the house | 92.1 (84.7–99.6) | 85.3 (74.0–96.6) | 76.5 (57.9–95.2) | 78.2 (64.1–92.4) | 1.10 (0.88–1.39) |
| Animal excrement on the house floor | 66.7 (55.8–77.6) | 65.1 (54.7–75.6) | 51.8 (45.8–57.9) | 57.4 (51.4–63.4) | 1.13 (0.84–1.54) |

**Table 2: Change in WASH-indicators in all households where a household survey was taken (A) and households where a household survey and stool was sampled (B).**
| B. Households with stool collection (n = 265) |  |  |  |  |  |
| --- | --- | --- | --- | --- | --- |
| Practice/condition | Baseline |  | Post-intervention |  | Adjusted PR |
|  | Control | Intervention | Control | Intervention |  |
| Use of unimproved drinking water source (dry season) | 15.3 (1.2–29.3) | 4.7 (0.0–10.0) | 13.2 (0.0–26.5) | 5.7 (1.7–9.8) | 1.42 (0.55–3.65) |
| Use of unimproved drinking water (rainy season) | 17.7 (2.2–33.1) | 8.3 (1.7–14.9) | 10.7 (1.4–19.9) | 4.9 (0.0–10.0) | 0.98 (0.37–2.63) |
| Incorrect handwashing** | 90.0 (79.8–100.3) | 83.0 (72.7–93.3) | 86.9 (75.4–98.4) | 86.1 (76.2–96.1) | 1.08 (0.90–1.29) |
| Using unimproved sanitary facility | 68.5 (53.8–83.2) | 67.7 (49.5–85.9) | 58.4 (38.7–78.0) | 62.1 (46.7–77.4) | 1.08 (0.71–1.63) |
| Livestock animals access the house | 87.0 (75.0–99.0) | 85.3 (72.2–98.4) | 71.9 (50.8–93.0) | 75.7 (60.7–90.6) | 1.07 (0.83–1.38) |
| Animal excrement on the house floor | 64.0 (55.9–72.2) | 63.3 (51.6–75.0) | 49.0 (41.7–56.3) | 51.5 (43.9–59.2) | 1.06 (0.73–1.56) |

PRs were consistently close to 1, i.e. 1·05 (0·91–1·21) for correct handwashing, and 1·13 (0·84–1·54) for animal excrement on household floors. Livestock access to household structures also remained high in both arms, with a PR of 1·10 (0·88–1·39).

## Discussion

Our pre-intervention estimates indicated a high burden of ESBL-E transmission at the household level in rural Burkina Faso. Pre-intervention colonisation prevalence was 56.6% in control villages and 47.2% in intervention villages, with corresponding incidence rates of 3.8 and 3.5 acquisitions per 100 person-days. This equates to approximately 1.1 and 1.0 acquisitions per household member per month, respectively, and nearly double that rate when averaged annually. These acquisition rates are around five times higher than those observed in Dutch households and substantially exceed estimates from other high-income household settings.^9,11^ Although not conclusive, our findings suggest that a community-level behavioural intervention targeting medicine providers and serving communities may have reduced ESBL-E acquisition 6–9 months after rollout (HR 0.82, 95% CrI: 0.56–1.14). The posterior distribution indicated an 88% probability of a reduction in acquisition hazard and a 71% probability of a reduction of a magnitude we considered meaningful. Taken together, these results are most likely consistent with a modest beneficial effect.

We did not identify measurable improvements in WASH behaviours in Nanoro following the intervention, however community-level dispensing of antibiotics declined substantially (adjusted risk ratio [aRR] 0·40; 95% CI 0·20–0·79).^21^ Given that antibiotic exposure can facilitate ESBL-E acquisition by suppressing susceptible gut flora, the observed reductions in antibiotic dispensing could have possibly contributed to the lower acquisition we measured.^15^ Delayed population-level responses to reduced antibiotic selection pressure have been documented elsewhere, with declines in *E. coli* resistance (including amoxicillin–clavulanate and ampicillin) lagging reductions in prescribing by 1–3 months.^24^ In Nanoro, relatively low baseline healthcare seeking, as well as low Watch and overall antibiotic dispensing (1·1 [95% CI 0·0–2·3] and 9·9 [95% CI 8·8–11·0] per 1000 inhabitants per month, respectively) may have constrained the magnitude of intervention effects.

However, ESBL-E acquisition risks almost doubled in the rainy season, suggesting that factors beyond antibiotic use contribute to community transmission. Ongoing work in Nanoro will further characterise seasonal patterns in antibiotic use,^25^ but prior work from the same, and another rural district in Burkina Faso (Nouna) found that antibiotic consumption among individuals attending primary health centres is actually lower during the rainy season, when malaria cases dominate clinical care.^26,27^ This finding points to environmental or ecological factors - in addition to antibiotic consumption - driving AMR fluctuations.^13,28^ Indeed, similar seasonal patterns have been documented in Malawi where ESBL-producing *E. coli* colonisation was associated with a range of environmental and household exposures, including drinking water from tube wells or boreholes, and proximity to open defecation.^29^ Potential mechanisms proposed in previous work include the accumulation of mud and floodwater, which may increase contact with contaminated soil and drinking water sources, including wells and boreholes.^22,29^ These patterns align with our findings, where unimproved sanitation - mainly open defecation - was associated with increased acquisition risk, and common use of boreholes. Moreover, recent genomic data from Kenya similarly show that households with safer water collection and storage exhibit reduced genetic clustering of ESBL-E, indicating lower within-household transmission.^14^ Improved drinking water in our setting did not involve chlorination at the point of use, which could explain why we did not observe similar associations with ESBL-E acquisition. Nonetheless, our findings combined with these, and other emerging large scale genomic evidence from similar settings,^30^ indicate that household- and community-level environmental exposures likely play a substantial role in sustaining high AMR transmission in rural low-resource settings. Consequently, high-AMR-burden rural settings may benefit from integrated One Health strategies that combine environmental risk reduction with antibiotic stewardship interventions, similar to our approach.^31^ Such interventions could prioritise the safety of drinking water sources (e.g. protection, maintenance, and point-of-use treatment of wells and boreholes), reducing environmental contamination through sanitation improvements, and strengthening hygiene and environmental control measures during the rainy season when transmission risk is highest.

At the same time, there remains a critical need for more robust, causal evidence on the relative contributions of community-level drivers of AMR, as well as on the (cost-)effective and sustainable delivery of WASH interventions in resource-poor settings. In view of this uncertainty, our intervention was intentionally implemented as a bundled strategy, which necessarily limits the extent to which observed effects can be attributed to specific components. A 2024 modelling analysis suggested that universal WASH alone could avert ∼247,800 (95% CI 160,000–337,800) AMR-associated deaths through reduced diarrhoea incidence each year, with further gains from infection prevention and control and vaccination.^32^ That said, one of the key lessons from the few large-scale randomised controlled WASH trials (which targeted childhood stunting and diarrhoea) is that sustaining individual-level WASH behaviours in rural, resource-limited settings is inherently difficult. This may explain the absence of measurable WASH behaviour change in our study and underscores the importance of addressing broader structural and environmental pathways involving engineered, infrastructural and systemic changes, alongside the need to co-design future solutions with communities.^33^

The trial was explicitly powered to detect intervention effects on ESBL-E acquisition and was supported by longitudinal microbiological sampling. While power calculations indicated a high probability of detecting a ≥30% reduction in acquisition, smaller effects such as those we observed may still be clinically relevant. To our knowledge, this is the first study in rural sSA to quantify household-level ESBL-E acquisition, a key precursor to clinical infection. By using continuous-time multi-state models, we were able to estimate acquisition dynamics and seasonal variation while accounting for staggered intervention timing and household-level heterogeneity, yielding statistically more efficient and mechanistically informative intervention endpoints than simple prevalence-based measures. Moreover, by estimating intervention effects conditional on individuals’ baseline colonisation status, baseline imbalances between intervention and control group prevalence were intrinsically controlled for. Hence, we provide a methodological framework for evaluating AMR and antibiotic stewardship interventions beyond current standard practice based solely on changes in targeted exposures. A limitation of our study was that our chosen microbiological sampling intervals did not allow for reliably distinguishing intervention effects on acquisition, from effects on decolonisation events. We opted for three and six months intervals, based on previously estimated mean duration of ESBL-E colonisation of three to four months,^34^ but methodological approaches to define optimal spacing could help future AMR intervention trials that aim to similarly assess microbiological impacts.^35^ Moreover, our approach does not account for baseline differences in characteristics that may influence transmission risk during follow-up, such as socioeconomic conditions, and residual confounding, potentially affecting exposure risk or intervention uptake, cannot be excluded. Furthermore, recent genomic analyses show that transmission rates can vary even within single *E. coli* clades, which we did not account for.^36^ However, as ESBL genes can spread across lineages and species via horizontal gene transfer^12^ we believe our estimates still capture the broader ARG transmission dynamics, relevant for estimating our intervention effect. Within CABU-EICO, we have sequenced more than 900 ESBL-E isolates, and ongoing whole-genome analyses will provide higher-resolution insight. By integrating these genomic data into a hidden Markov or multi-state modelling framework,^37^ we aim to further disentangle within-community transmission pathways and quantify their links to environmental exposures.^21^

### Conclusion

Our findings are consistent with a modest reduction in household-level ESBL-E incidence following a contextualised, community-based behavioural intervention targeting both medicine providers and communities in rural Burkina Faso, with effects emerging several months after intervention implementation. Further research disentangling the individual and combined effects of the intervention components, and identifying how these interact with the environmental and seasonal drivers of ESBL-E acquisition identified in this study, will be important for guiding more targeted One Health intervention strategies in Burkina Faso and, more broadly, across rural sSA.

## Data Availability

All data produced are available online at https://github.com/RaneemAizouk/CABU-EICO/tree/main/Scripts

https://github.com/RaneemAizouk/CABU-EICO

https://github.com/ingelbeen/cabu_intervention

## Acknowledgements

We are grateful for microbiological guidance provided by Prof Jan Jacobs. We are also very grateful to the participants and would like to acknowledge the effort provided by the drivers, administrative staff, field workers, lab staff, and community health workers. Specific thanks go to the members of the CABU-EICO study team from both the Nanoro and Kimpese study sites that have supported either the social sciences work that fed into the intervention design, study coordination, data collection, curation, and those that were involved in the remainder CABU EICO work packages on rodent AMR surveillance and molecular characterisation (alphabetical order): Iana Amke-Lowrey-Gold, César-Augustin Mambuku Khoso Muaka, Djibril Binga, Tamara Giles-Vernick, Leonard Heyerdahl, Oscar Kiabanza, Herwig Leirs, Adna Melanda, Richelin Makuaya, Joachim Marien, Didier Ndomba, Anicet Nzuzi, Delphin Mavinga Phanzu, Toussaint Rouamba, Franck Sovi Hien, Eric Wendpouiré Tiendrébeogo, Sandra Van Puyvelde, Rianne van Vredendaal.

## Author contributions

EvK, BSC, BI, DV, and MvdS conceptualised the study; LC, KJS, WA, and MM coordinated the intervention development and implementation. LC and DPM oversaw the photovoice activities - that informed the intervention design. BI, and DV developed and validated the data collection questionnaires, with input from MvdS and EvK; YS coordinated the microbiological analyses; BR developed the WASH coding framework for intervention evaluation. DV, and BI coordinated data collection; EvK curated, and validated the data; RA and EvK analysed the data, with input from BSC; EvK wrote the original draft manuscript with input from RA; all authors revised and edited the first and subsequent drafts of the manuscript; all authors had full access to the study data and had final responsibility for the decision to submit for publication.

## Data sharing

Pre-study sample size calculations are published at: https://github.com/esthervankleef/sample_size_jpiamr. Questionnaires, informed consent forms, pseudonymized datasets, data dictionaries, and the full analysis code is publicly available under an open-access license at: https://github.com/RaneemAizouk/CABU-EICO/tree/main. The study protocol was published: https://doi.org/10.1186/s13063-023-07856-2.

