## Supplementary material for "Targeting community-level drivers of antimicrobial resistance in sub-Saharan Africa: the effect of a community-based intervention bundle on household transmission of Extended Spectrum Beta-lactamase-producing *E. coli* in rural Burkina Faso - a cluster randomised trial"

**The effect of a community-based intervention bundle on household transmission of Extended Spectrum Beta-lactamase-producing *E. coli* in rural Burkina Faso - a cluster randomised trial - Supplementary material**
 *[this work is now accepted for publication in Lancet Microbe]*

### Section 1: Study design

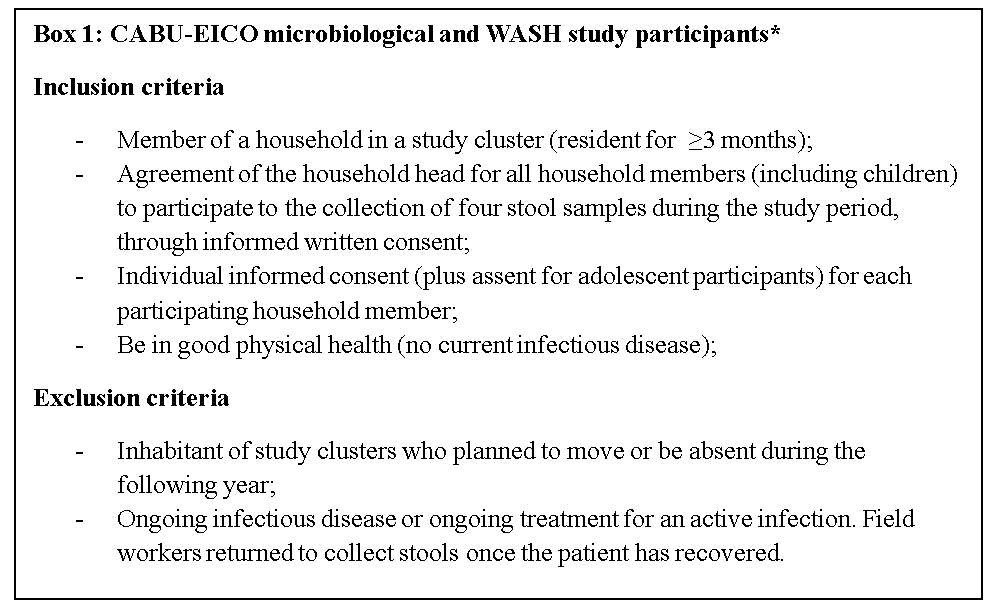

### **Box 1: CABU-EICO household survey in- and exclusion criteria**. *For the households where only the WASH survey was conducted and not stool sampling, the household head signed informed consent

### Section 2: Data collection surveys

HOUSEHOLD, HEALTHCARE SEEKING AND STOOL COLLECTION SURVEY

This document consists of two questionnaires:

The household survey on WASH behaviour and healthcare seeking behaviour (Household-level, Round 0, Oct-Dec 2022 and Round 3, Oct-Dec 2023)

1) The household stool collection survey for households selected for stool collection (Individual-level, R0 Oct-Dec 2022, R1, R2, R3)

Questionnaire on WASH practices and healthcare utilisation
(one per household)

General information

Household ID |__|__|__|

Survey date (dd/mmm/yy)

Initials of head of household or person answering questions |__|__|__|

Number of people in the household |__|__|

Number of children aged 0 to 59 months (0 to under 5 years)

Number of households in concession |__|__|

Water, sanitation and hygiene

**Knowledge**

1. Diarrhoea can be prevented by which of the following measures?

|__| Keep the water pot covered in the house

|__| Do not dip your fingers in a glass of drinking water

|__| Use a utensil with a handle to take the water from the pot.

|__| Cover food

|__| Boil drinking water

|__| Filter drinking water

|__| Other

|__| Can't be avoided

|__| Don't know

**Drinking water**

2. What is your main source of drinking water during the DRY season?

|__| Faucet in the household

|__| Faucet in the concession

|__| Public tap/water fountain

|__| Borehole

|__| Well (protected from external contamination)

|__| Unimproved well (unprotected)

|__| Rainwater

|__| Surface water (ponds, dams, rivers, lakes, pits, irrigation canals)

|__| Water in sachets

|__ |Bottled water

3. What is your main source of drinking water during the RAINY season?

|__| Faucet in the household

|__| Faucet in the concession

|__| Public tap/water fountain

|__| Borehole

|__| Well (protected from external contamination)

|__| Unimproved well (unprotected)

|__| Rainwater

|__| Surface water (ponds, dams, rivers, lakes, pits, irrigation canals)

|__| Water in sachets

|__ |Bottled water

4. Are cans (including a large tank) used to store drinking water in the house?

|__| Yes, cans

|__| Yes, only one large tank

|__| No

5. If yes,
   a. Are the cans (or tank) closed when filled with water? Y/N
   b. Are the cans (or tank) closed when empty? Y/N
   c. Cans (or tanks) are cleaned before being reused or refilled?

|__|Yes, with soap (or washing-up liquid) and water.

d. |__|yes, with water but no soap
e. |__| yes, boiled
f. |__| no

6. Do you treat your drinking water?

|__| We don't treat it

|__| Boil

|__| Chlorinate/add disinfectant

|__| Filter with cloth

|__| Filter with a filter

|__| Solar disinfection (do you leave the water in the sun for a while?)

|__| Decant

|__| Other, please specify ___________

**Defecation**

7. Which type of sanitation facility is the most important for your household? (one option only)

|__| Flush toilets connected to a septic tank

|__| Improved pit latrine with ventilation

|__| Pit latrine with slab

|__| Pit latrine without slab

|__| open defecation

|__| Other, specify ..........

8. What other types of defecation do members of your household perform when they are at home (at least once a week on average over the year)? (several options possible)

|__| Flush toilets connected to a septic tank

|__| Improved pit latrine with ventilation

|__| Pit latrine with slab

|__| Pit latrine without slab

|__| open defecation

|__| Other, specify ..........

9. If a toilet/latrine is used, is it shared with other households?

|__| Yes, other households known (non-public)

|__| Yes, shared with the general public

|__| No

10. If the answer was "yes", shared with how many other households? |__|__|
11. When was the last time the toilet or latrine was cleaned (if the respondent does not know because another household takes care of it, think about how often it is usually cleaned)?

|__| Within the last 24 hours

|__| >24 hours but < one week ago

|__| 1 to 4 weeks ago

|__| > one month ago

|__| Never

|__| Don't know

12. If there is a child under the age of two in the household: The last time [child's name] had a bowel movement, how was the stool disposed of?

|__| The child has used the toilet/latrine

|__| Thrown/rinsed into toilet or latrine

|__| Thrown/rinsed into a drain or ditch

|__| Thrown in the garbage (solid waste)

|__| Buried

|__| Disposed of in the open air

|__| Used as manure

|__| Other (specify)

|__| Not applicable (no children <2 years old)

13. If toilet, latrine, or pit: the last time the (septic) tank was emptied, where were the contents disposed of?

|__| Has not yet been drained

|__| Don't know

|__|Removed by the service provider to a treatment plant

|__| Removed by the service provider and buried in a covered pit

|__| Removed by service provider (don't know where)

|__|Emptied by household and buried in covered pit

|__|Emptied by household into uncovered pit, open ground, body of water or elsewhere

|__|Other (specify)

**Hand washing**

14. Is there any product (soap, detergent, ash) for washing hands in use (wet, clearly used in the last 24 hours) in your household (survey observation)?

|__| Yes, bar or liquid soap

|__| Yes, detergent used (powder/liquid/paste)

|__| Yes, ash/mud/sand used

|__| No, no washing product available

|__| No, a washing product available but not used in the last 24 hours

15. If yes, do you always wash your hands with soap after defecation? (check by observation: is the hand-washing point so close to the toilet that this is plausible?)

|__| Yes, always

|__| Yes, often but not always

|__| No or rarely

|__| Not sure

16. If yes, do you always wash your hands with soap before meals (check by observation: is the hand-washing point so close to the eating area that it is plausible)?

|__| Yes, always

|__| Yes, often but not always

|__| No or rarely

|__| Not sure

**Animal contact**

17. Do you keep any animals (oxen, sheep, goats, pigs, donkeys, horses, chickens, geese, ducks) on the plot where the household lives?

|__| Yes, inside and outside the house

|__| Yes, outside but adjoining the house

|__| Yes, outside but within a defined area

|__| No

18. If "Yes, inside and outside the house", tick the animals in the house (several options possible)

|__| Oxen

|__| Sheep or goats

|__| Pigs

|__| Donkeys, horses

|__| Chickens, geese or ducks

|__| Other, Specify ________

19. Which animals are kept out of the house (several options possible)

|__| Oxen

|__| Sheep or goats

|__| Pigs

|__| Donkeys, horses

|__| Chickens, geese or ducks

|__| Other, Specify ________

20. Observation: Animal excrement on the floor of the house

|__| yes

|__| no

|__| not possible to determine

21. Have you given antibiotics to the animals (check the medication or prescription if available, and compare against the provided list of antibiotics)?
    |__| Yes, definitely an antibiotic
    |__| Possibly an antibiotic (a medicine was given, but it’s not certain whether it was an antibiotic or it’s not present in the household)
    |__| No
    |__| Don’t know
22. If the answer was “Yes, definitely an antibiotic,”
    a. where was this/these antibiotic(s) purchased? (specify whether it was from a drug depot or formal pharmacy, an informal medicine seller, or a veterinarian) (text response)
    b. Specify the antibiotic (generic name, route of administration, dosage, regimen, number of units) (text response)
    c. For which animal was the antibiotic used?
23. When an animal on the plot is sick, what do you do with it (several options possible)?

|__| Bring in a veterinarian who can prescribe a treatment

|__| Buy treatment without veterinary consultation

|__| Sell the animal to a butcher

|__| Eat the meat at home

|__| When the animal dies, throw it away or bury it

Foodborne risk

24. Do any household members regularly drink (at least once a week) raw/fresh milk without boiling or pasteurizing? (multiple options possible)
    |__| Yes, cow’s milk
    |__| Yes, goat’s milk
    |__| No

Frequency/choice of care-seeking
(one per household)

1. Do you currently have any medicines that you keep in your household that you use often when you need them?

|__| Yes

|__| No

2. If so, can you show them to us?
3. Antibiotics (or medicines that could be antibiotics): Specify the generic name, method of administration and dosage (in mg, or if suspended the concentration, e.g. 200mg/5mL), and number of units (tablets, bottles, ampoules).
   a. Medicine 1 ……………………………………………………….
   b. Medicine 2 …………………………………………………………
   c. Medicine 3 ………………………………………………………….
   d. Medicine 4 ………………………………………………………….
   e. Medicine 5 ………………………………………………………….
4. If you're not sure whether the medicine is an antibiotic, take a photo
5. DURING THE LAST THREE MONTHS, for all the members of your household, how many times did you go to seek health care (including visits to pharmacies or other medicine sellers)?

__ (integer)

6. DURING THE LAST THREE MONTHS, how often did all the members of your household use medicines stored at home or not obtained from a medicine suppliers? If a member visited more than one health facility or medicine provider, please enter one loop/repeat per visit. For example: If a member of the household was ill twice and visited two providers for the first illness, and took medicines stored at home for the second, without visiting a dispenser, three episodes of care-seeking should be entered.

|__|

___________

From here, a loop (repeat) will be entered in ODK to individually document each episode of care-seeking by each member of the household, including self-medication. If a member visited several health facilities or drug providers, please enter one loop/repeat per visit.

________________

1. ID of household member (e.g. m1, m2, etc.) ________
2. age (in years) of this household member (if less than two years, convert months to years with two decimals. e.g. 3 months = 0.25) ___
3. Date of visit or use of medication
   a. in the week prior to this survey
   b. more than a week but less than a month before this survey
   c. between one and three months prior to this survey
4. type of health care provider or drug seller (one choice on
   a. |__|With medicines we have at home
   b. |__|From medicine sellers at the market?
   c. |__| From traditional healers
   d. |__|Directly from a private pharmacy
   e. |__|Directly from the private depot
   f. |__|Directly from the public depot (inside health centres)
   g. |__|We first consulted the health centre (CSPS, CM, CMA) and bought the medicines as prescribed by the health worker.
5. If the answer was "medicines we have at home", specify the antibiotics used during this episode?
   a. |__| Amoxicillin oral
   b. |__| Amoxicillin + clavulanic acid oral
   c. |__| Phenoxymethylpenicillin (Peni V) oral
   d. |__| Metronidazole oral
   e. |__| Cotrimoxazole (sulfamethoxazole + trimethoprim) oral
   f. |__| Erythromycin oral
   g. |__| Azithromycin oral
   h. |__| Ciprofloxacin oral
   i. |__| Ampicillin parenteral
   j. |__| Gentamycin parenteral
   k. |__| Ceftriaxone parenteral
   l. |__| Other, specify .........................
   m. |__| None

6. Symptoms presented (several options possible)

a. |__| Fever
b. |__| Cough
c. |__| Cold
d. |__| Sore Throat
e. |__| Headache
f. |__| Diarrhea
g. |__| Vomiting
h. |__| Sores
i. |__| Fatigue
j. |__| Burning when urinating
k. |__| Other, please specify ...........................

7. time in days between onset of first signs and visit or use of medication ___ (even if not exact, try to make an estimate, so that you can understand the sequence of visits to different providers for the same episode of illness)

—-End of loop—---

End of household questionnaire.

If this household is one of those with stool collection, please complete a stool collection form for each member of the household.

Stool collection form
(one form per individual collected)

*Complete one form per household member to collect a stool sample.*

Individual ID of household member |__|__|__|

This member of the household:

|__|Was present and gave consent (or parental consent was given)

|__|Was present but did not give consent (or parental consent)

|__|Was absent and a copy of the consent form will be left in the household

Date of consent given (if "present and gave consent", if "absent", to be entered the following day if consent was given) (dd/mm/yy)

Sample number (label on container) |__|__|__|

If the date for collecting the container with the sample is not the next working day, please specify when the sample should be collected. (text answer)

*______*

*To be completed after stool collection*

Time of stool production/defecation _ _ h _ _

Time samples collected _ _ h _ _

**End of questionnaire**

### Section 3: Statistical analyses

#### *Power calculations for ESBL-E household transmission outcome*

We assumed a continuous-time Markov model of household transmission with two states (colonised/ not colonised), with transitions governed by acquisition (transmission) and decolonisation rates. Parameters were informed by a Dutch study,^10^ i.e. the same decolonisation rate, a background transmission rate ten times higher, and a pre-intervention person-to-person within-household transmission rate twice the rate estimated in the Netherlands. Synthetic datasets (N=1000 per intervention effect scenario) were generated under these assumptions for households of size 5, with 132 households per arm and observation times at 0.25, 0.5, and 1 year. These design choices were informed by laboratory capacity at the Nanoro site. Each simulated dataset was analysed with a generalised estimating equation approach (R package *gee*) accounting for household clustering with a Poisson family and an AR-1 correlation structure using robust standard errors. Empirical power was estimated as the proportion of replicates with two-sided p<0.05 for the intervention effect on acquisition coefficient; 95% CIs were computed by binomial approximation.. Assuming the intervention reduced the transmission rate by 40%, 30%, and 20% respectively the study was then estimated to have 99%, 82% and 43% power respectively to detect a reduction in transmission. This informed the choice of 12 randomly selected HDSS households per cluster (132 households per intervention arm). R code can be found here <https://github.com/esthervankleef/sample_size_jpiamr>. The data-generating model used for these simulations represents a simplified approximation of the primary analysis model and does not incorporate additional features such as seasonality. Code is available at: <https://github.com/esthervankleef/sample_size_jpiamr>. Of note: simulation-based model checking (see corresponding section) was performed using data generated from the final analysis model.

#### *Continuous-Time Multi-State Modelling Framework*

We used a Bayesian continuous-time markov chain (CTMC) modelling framework to estimate the transitions between colonisation states over time. This stochastic process assumes that, at any given time, the system is in one of two states, i.e. *S₁*: uncolonised with ESBL-E and *S₂*: colonised with detectable ESBL-E. Transitions are allowed in both directions, where *S₁*→*S₂* represents acquisition and *S₂→S₁* decolonisation. We denoted the instantaneous acquisition hazard rate at time *t* as *λ_12_(t)* and the instantaneous decolonisation hazard rate at time *t* as *λ_21_(t).* Conditional on being in state S_i_ at time *t*, the waiting time *x* until a transition to state *S_j_* occurs is exponentially distributed with rate *λ_ij_(t).* For a given piecewise-constant rate *λ* > 0, the probability density function of the waiting time distribution is

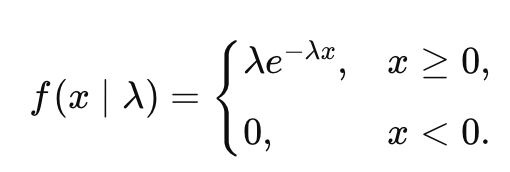
(Eq. 1)

This formulation implies the Markov property, whereby the future state of an individual depends only on their current state and the associated transition intensities. At any time (*t*), the CTMC is defined by the transition intensity matrix:

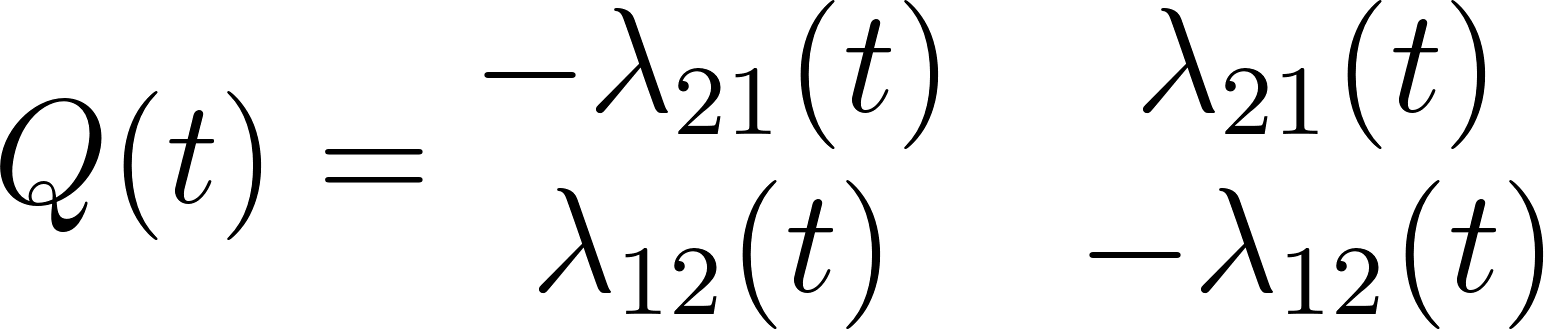
(Eq. 2)

Transition rates λ_12_(t) and λ_21_(t) were modelled as functions of individual- and household-level factors as well as time-varying environmental effects. Specifically, the CTMC incorporated individual covariates (age, sex), household-level random effects *µ_h_* to account for clustering, and seasonal variation *s(t)*.

##### *Seasonal terms*

To allow for capturing seasonal temporal trends in acquisition ESBL-E (ESBL-E colonisation times were not assumed to be subject to seasonal patterns, see later), as well as time-varying intervention effects, the overall study period was subdivided into 28-day intervals (~one month). Within each 28-day subinterval, hazards were assumed constant, and both seasonal and intervention timings were evaluated at the midpoint of the subinterval. During model development, we examined monthly acquisition and prevalence trends stratified by intervention arm to assess potential season-by-intervention interaction. As temporal patterns were similar between groups (Figure S4), no interaction term was included in the final model. Then, for a time interval of length $\Delta t$ (generally 28 days), the corresponding transition probability matrix is:

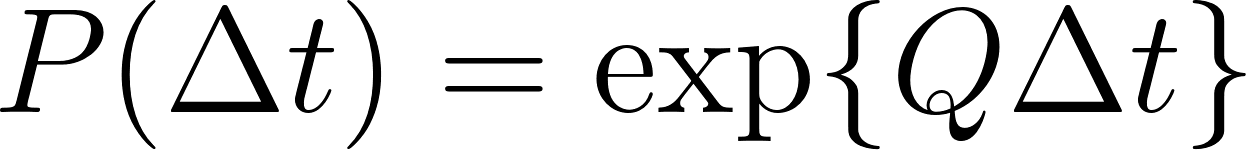

(Eq. 3)

This matrix gives the probability of being colonised or uncolonised at the end of the subinterval, conditional on the state at its start. In the two-state CTMC, each element of these transition probability matrices can be expressed in closed form as

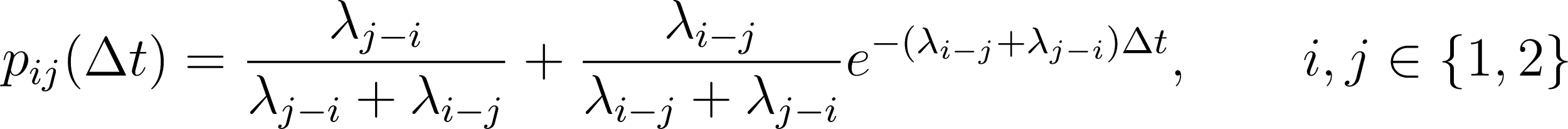

(Eq. 4)

where *p_ij_* represents the probability of transitioning from state i to state j over an interval of length Δt. This simplifies to

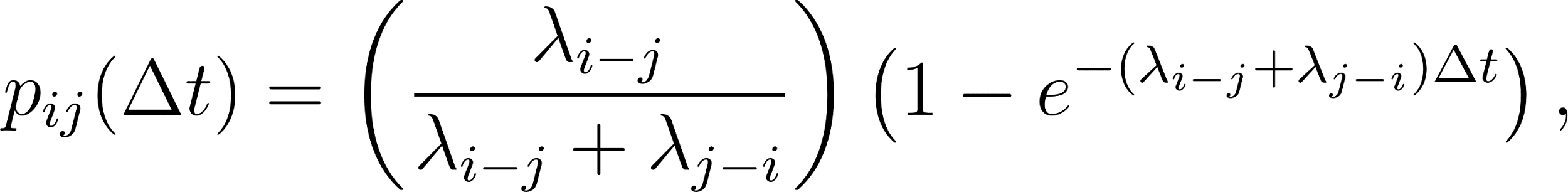

(Eq. 5)

and by complementarity

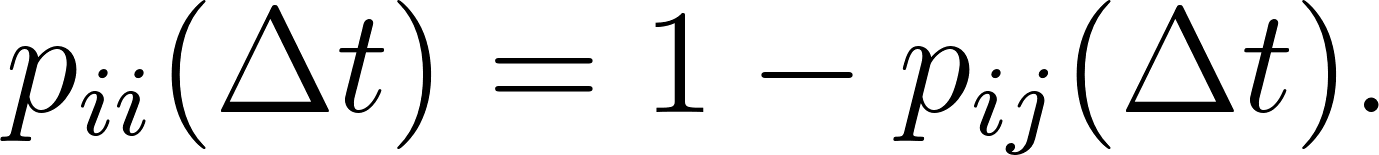

(Eq. 6)

Each 28-day interval thus produced an interval specific transition probability matrix

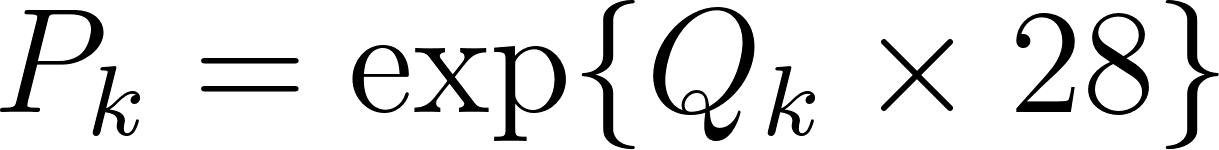

(Eq. 7)

For each period between sampling times the overall transition probability was obtained by multiplying the relevant transition matrices:
(Eq. 8)
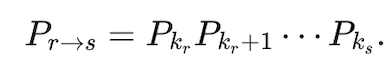

We modelled seasonality only for the acquisition rate *λ*_12_(t) as environmental (rain vs dry season-related) exposures were assumed to alter risk of acquiring ESBL-E but not expected to affect colonisation times. To capture seasonal variation in transition rates, we evaluated two alternative formulations for the seasonal component *s(t)*, i.e. a sinusoidal annual cycle (parametric), according to:

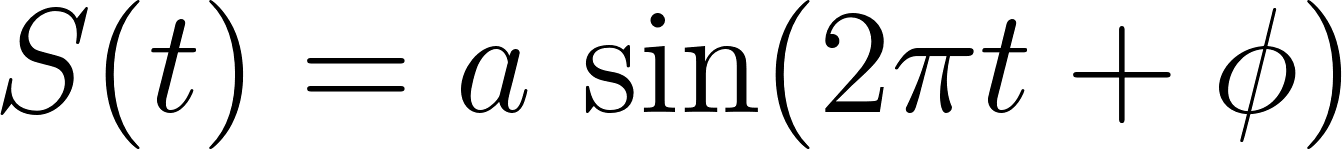

(Eq. 9)

where $a$ represents the amplitude, governing the strength of seasonal variation; and *Φ* is the phase shift, determining where the peak of the seasonal cycle occurs in the calendar year. This formulation assumes a smooth, regular seasonal oscillation with one cycle per year. Secondly, we fitted models with a periodic B-spline representation (non-parameter, i.e. more flexible), i.e:

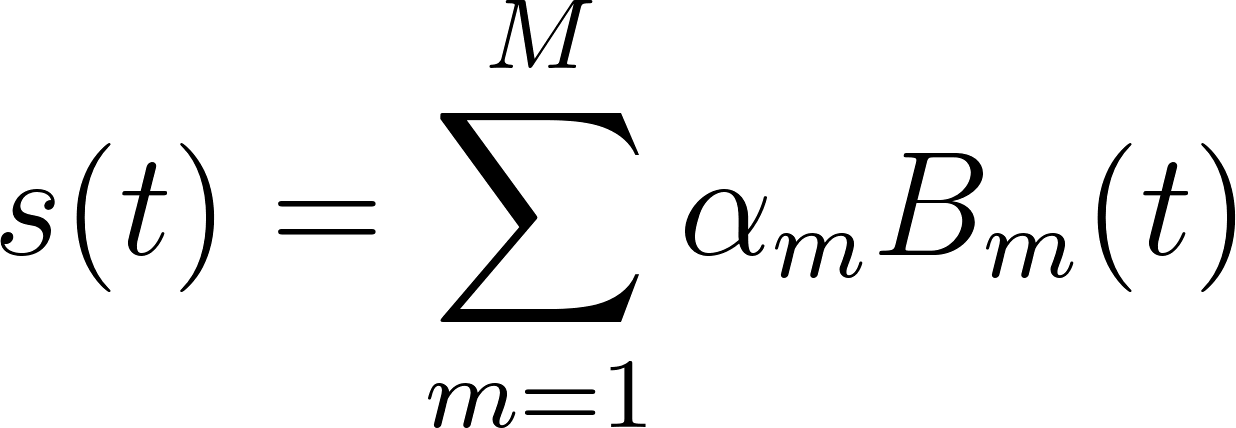

(Eq. 10)

where *B*_m_(t) denotes periodic spline basis functions defined over the calendar year. The coefficients *α*_m_ determine the amplitude and shape contributed by each basis function. This formulation provides a flexible, non-parametric representation of seasonality, allowing the data to determine the shape of the annual pattern.

##### *Intervention terms*

The intervention bundle was delivered in three rounds. In our base case scenario, we grouped intervention rounds 1 and 2 into a single early intervention effect *β*_intv1_ and treated round 3 as an additional late intervention effect *β*_intv2_. For each village we specified two calendar times, *t*_intv1_ and *t*_intv2_, marking completion of the early (round 1 and round 2) and late (round 3) intervention rounds. In the likelihood, each observation interval was split wherever it crossed these village-specific intervention dates, so that segments contributed person-time as (i) pre-intervention, (ii) exposed to the early intervention only, or (iii) exposed to both early and late intervention. Correspondingly, in the basecase scenario, the log-hazards for acquisition included two time-varying terms, *β*_intv1_ and *β*_intv2_, which modified the baseline, age, sex, seasonal and household effects once *t* ≥ *t*_intv1_ and *t* ≥ *t*_intv2_, respectively according to

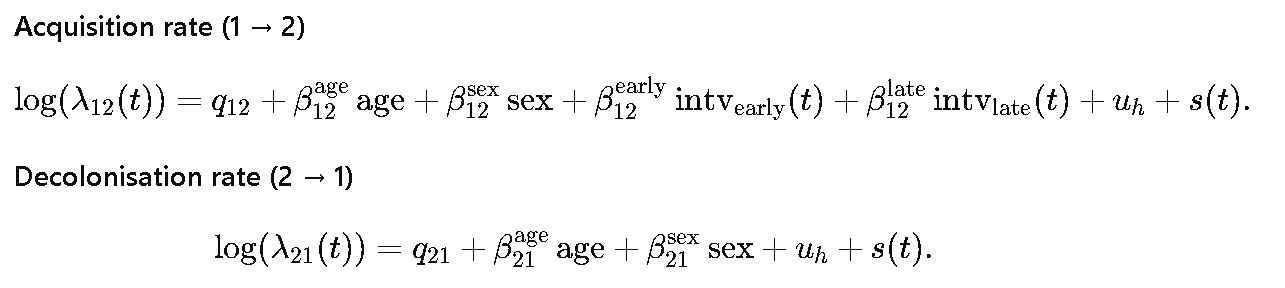

(Eq. 11)

while decolonisation log-hazards were estimated according to

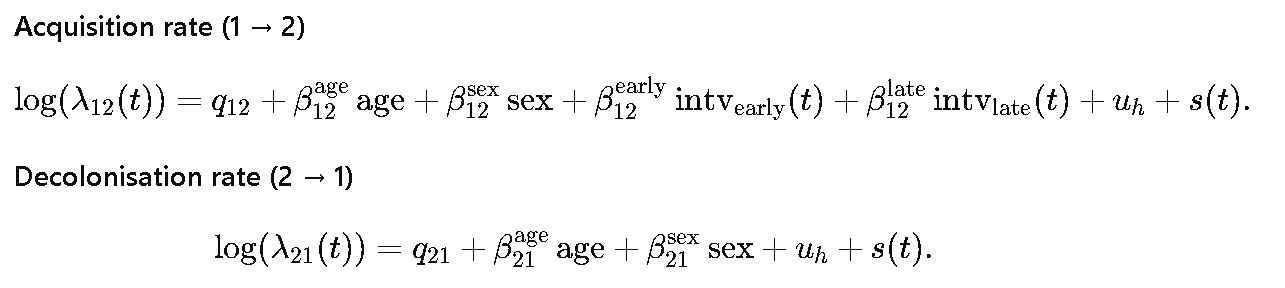

(Eq. 12)

*Estimation of intervention effects - hazard and incidence ratios*

We subsequently quantified the effect of the intervention using two complementary measures: (i) hazard ratios (HRs), reflecting instantaneous effects on the acquisition hazard, and (ii) incidence rate ratios (IRRs), reflecting realised population-level differences in acquisition over the full follow-up period. For (i) we derived the hazard ratios for the intervention effect *β*_intv1_ and *β*_intv2_ as a measure for our early and late intervention effects respectively. Posterior samples of these coefficients were exponentiated to obtain hazard ratios (HRs). For (ii) we computed acquisition incidence per 100 person-days by aggregating interval-specific hazards according to the duration of each follow-up interval. For each posterior draw *d*, expected events (acquisitions) were calculated as *λ_12_(t)* × duration (dur) of each subinterval *r* and summed across intervals; dividing by total follow-up time in each phase *k* (early and late) yielded a time-weighted incidence rate rather than a simple mean hazard per intervention group *g* (intervention and control).

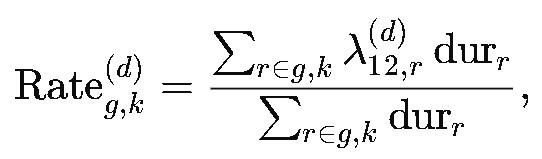

(Eq. 13)

Incidence rate ratios (IRRs) were then computed per posterior draw and phase *k* as a ratio-of-ratios (difference-in-differences on the multiplicative scale):

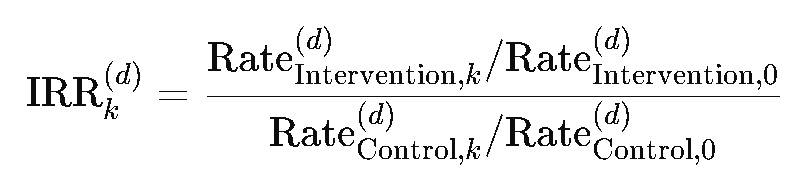

(Eq. 14)

k=0 denotes the baseline period. This captures the relative change from baseline in the intervention group compared with the corresponding change in the control group. Of note, because intervention and control groups could contribute follow-up in different calendar periods, control incidence estimates were standardised to the same calendar-time distribution as the intervention group. This ensured that differences reflect intervention effects rather than seasonal differences in underlying risk. This procedure was also repeated for the posterior draws from each of the village clusters separately.

##### *Prior specification*

We used weakly informative priors, but centred the baseline acquisition and decolonisation hazards on biologically plausible values informed by literature. The baseline log-acquisition rate was centred at −4.547 corresponding to an assumed acquisition rate approximately two-times that reported in high-income community settings, while the baseline log-decolonisation rate was centred at −5.154, consistent with a median colonisation duration of approximately four months.^10^ Deviations from these baseline values were allowed for, using standardised ‘raw’ parameters, with

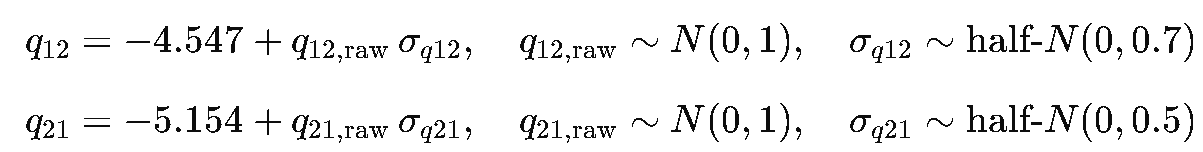

Covariate effects for age and sex on both hazards were assigned *N*(0,1) priors. Intervention effects (early and late) modified only the acquisition hazard in scenarios 1 and 2, with *β*_intv1 ,_ *β*_intv2_∼*N*(0,1) and no intervention effect on decolonisation, which was relaxed for scenario 3 and 4, using similar priors. Household-level random effects were modelled as

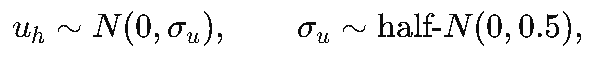

allowing moderate between-household variation. For models including sinusoidal seasonality, the amplitude parameter was given a *N*(0.5,1) prior, and the phase parameter a constrained *N*(π/2,0.5) prior on [0,2π]. For spline-based seasonality, each spline coefficient received *N*(0,1) priors. These priors discourage implausible hazard values while remaining sufficiently broad given uncertainty in ESBL-E transmission dynamics in community settings.

##### *Model fitting*

Models were estimated by Markov chain Monte Carlo using Stan’s No-U-Turn Sampler (NUTS), a variant of Hamiltonian Monte Carlo (HMC) with 4 chains, 5000 iterations per chain and 2000 warm-up iterations. We assessed model convergence by inspecting chain traceplots, R-hat values (~1) and absence of divergent transitions. We assessed comparative model fit of models with and without terms for seasonality and different intervention impact scenarios using leave-one-out cross-validation (LOO-CV). All models were implemented in *Stan version 2.23.2*.

##### *Simulation-based model checking*

We conducted simulation-based checks to evaluate whether the modelling framework could reproduce key features of the data and recover known underlying parameters. Using Stan’s *fixed_param* mode, synthetic datasets were generated from the final model structure using pre-specified values for intervention effects, participant-level covariates, and seasonal variation. Seasonal patterns were investigated via a sine-wave function over calendar time, with additional datasets generated in which we excluded the seasonality for comparison purposes. These simulated datasets were then analysed using models specifying seasonality either as a cubic B-spline or a sine-wave, allowing assessment of whether the true seasonal structure was recoverable. In the generated quantities block, latent states were tracked, and acquisition and decolonisation events were tallied across subintervals. Posterior predictive summaries including prevalence trajectories and acquisition counts were compared with the corresponding simulated values to evaluate how well the model reproduced the imposed dynamics.

To assess identifiability, we performed a parameter recovery analysis in which data were simulated from a known set of “true’’ parameter values under the same hierarchical and time-varying structure. The model was refitted to these synthetic datasets, and posterior estimates were compared with the parameters used to generate the data. The close agreement between estimated and true values indicated good parameter recovery, supporting model identifiability and internal validity.

##### *Prior and posterior predictive checks*

We performed a prior predictive check to determine the implications of our prior assumptions. Parameters were sampled from the prior distributions, and we used those to simulate the data using our Markov model, considering the time period of our study. The behavior of the simulated datasets was evaluated to verify that the priors generated outcomes within a plausible range. This evaluation confirmed the appropriateness of the prior specification before model fitting with the observed data. We performed posterior predictive checks to evaluate how well the fitted models reproduced key features of the observed data (Figure S1). Using draws from the posterior distribution, we generated replicated datasets under the model and summarised the predicted number of colonised individuals by study round and trial arm. These posterior predictions were then compared with the corresponding observed totals. We assessed whether the observed counts fell within the model’s 95% posterior predictive intervals for all three rounds and both arms, indicating that the fitted models were able to reproduce the observed data.

### Section 4: Supplementary figures and tables

#### *Supplementary figures*

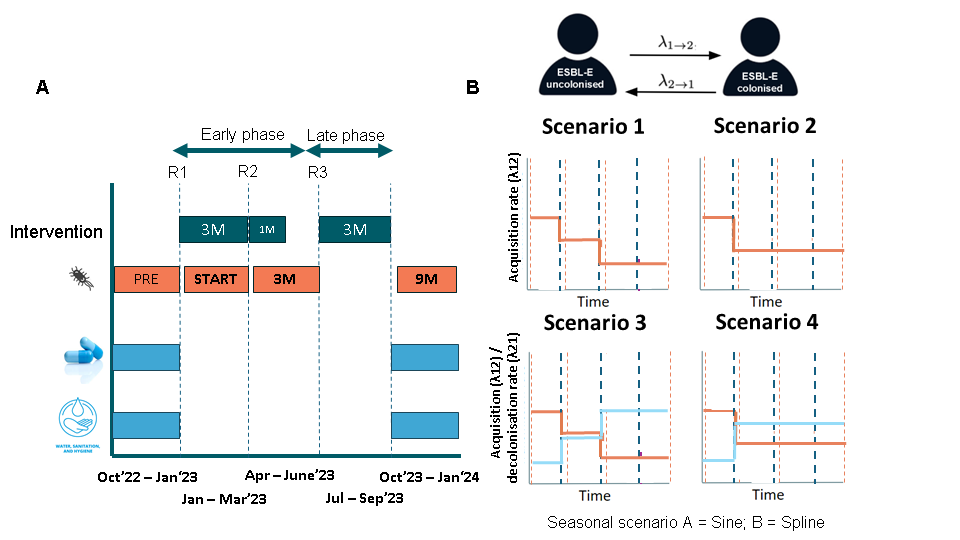

**Figure S1: Intervention and study design and data collection scheme (A); intervention effect scenarios considered (B); Modelling framework (C). A)** R1–R3 denote the three intervention rounds. Sea-blue dotted lines mark the start of each of the three intervention rounds. Coral dotted lines mark the start of each of the stool collection rounds, which were planned one week after the roll-out of each of the intervention rounds. The interventions were implemented in each of the 11 villages consecutively, spanning for round one and round three a three-month period. Round two was implemented over one-month to ensure the activities were implemented before the rainy season. Each of the stool collection rounds were anticipated to start one week after the introduction of the intervention in each of the respective village clusters, again over a three month period. **B)** λ₁₂ corresponds to the instantaneous daily rate of ESBL-E acquisition, whereas λ₂₁ represents the daily decolonisation rate. C) Scenarios 1–4 represent different assumptions about the timing and persistence of the intervention effect (e.g. no effect, immediate effect, delayed effect, and sustained or time-varying effects). Seasonal scenarios A and B represent alternative assumptions regarding the presence and form of seasonal variation in ESBL-E acquisition (e.g. absence vs inclusion of seasonal effects).

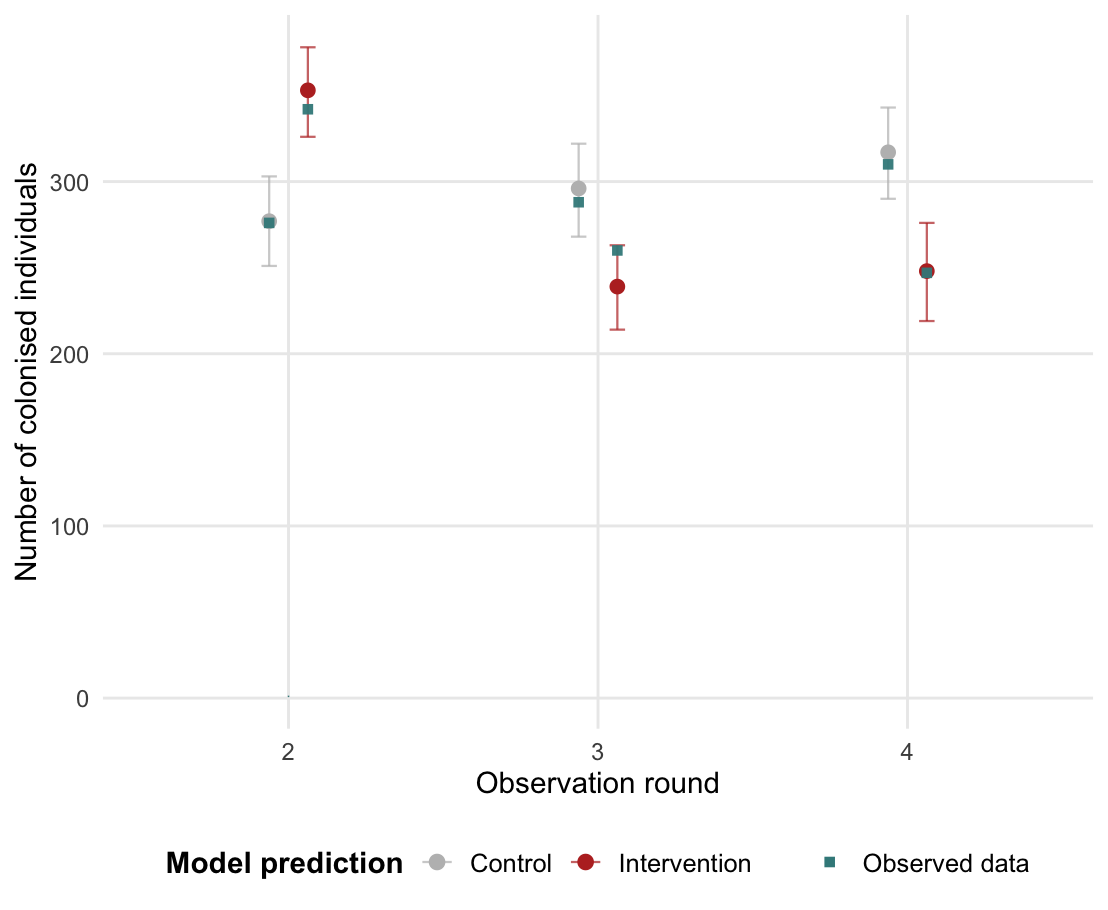

**Figure S 2: Posterior predictive assessment of base case scenario 1 model results.** Points show posterior predictive estimates of the number of colonised individuals for control (grey) and intervention (red) clusters across observation rounds 2–4; error bars represent the 95% credible intervals. Teal points represent the observed data. The close alignment between predicted and observed values across rounds indicates that the model adequately reproduces the overall patterns seen in the study population.

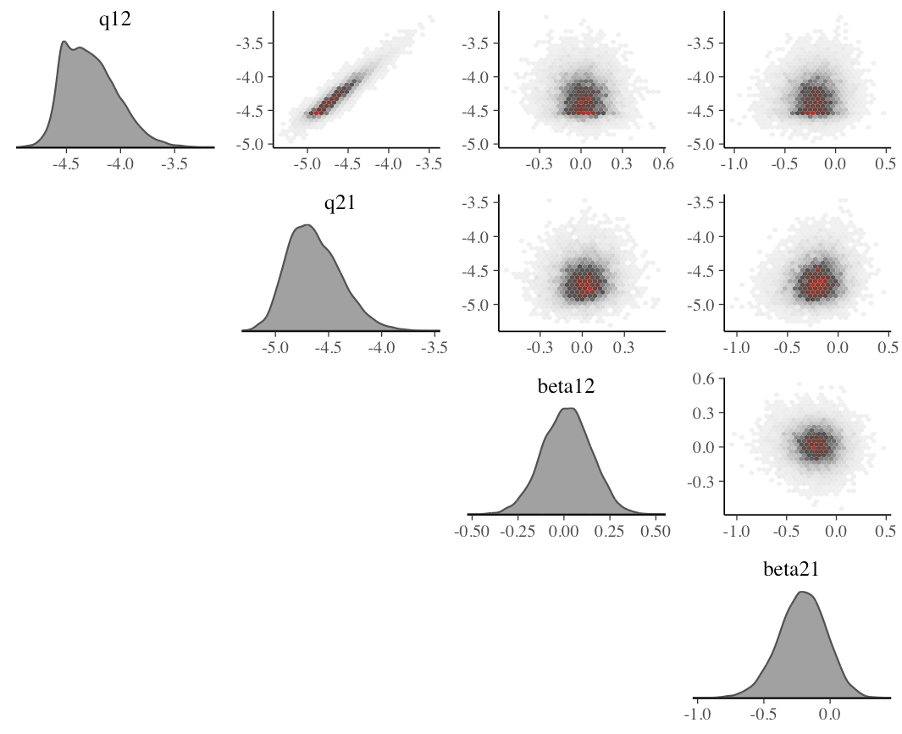

**Figure S 3: Posterior correlation structure between acquisition and decolonisation model parameter estimates.** q12: Baseline acquisition rate (transition from uncolonised to colonised); q21 baseline decolonisation rate (transition from ESBL-E colonised to uncolonised); beta12 intervention effect on acquisition rate; beta21: intervention effect on decolonisation rate. Shading reflects posterior density: light grey indicates low posterior density, dark red indicates high posterior density.

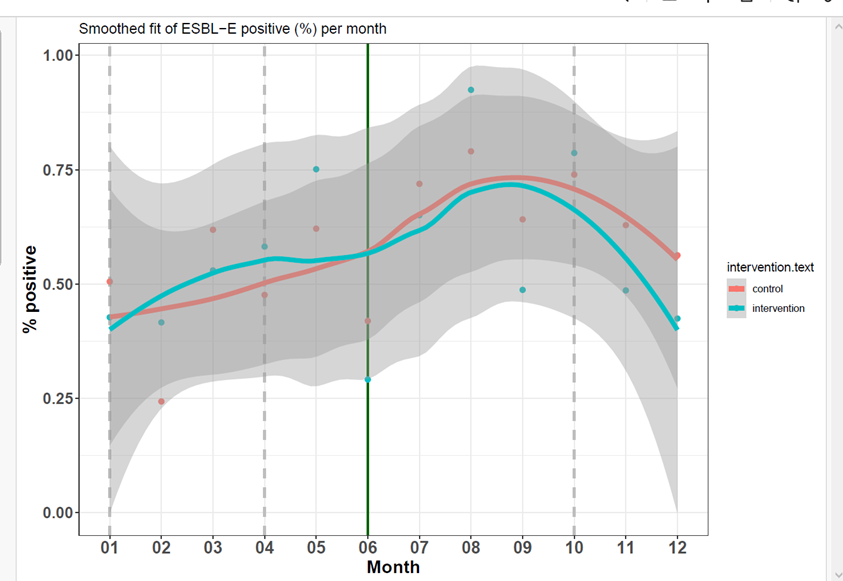

**Figure S4: Monthly ESBL-E prevalence stratified by intervention arm**. Points represent observed monthly proportions. Solid lines show locally weighted regression (LOESS) smooths fitted using the default geom_smooth() method in ggplot2, with shaded areas indicating 95% confidence intervals. Temporal patterns were similar in the intervention and control groups, with peaks occurring during the same months.

#### **Supplementary tables**

| Intervention component | Topic |
| --- | --- |
| **Face-to-face educational sessions (enabling) with healthcare providers and medicine dispensers** | |
| - clinical case discussion (all providers), prescription audit and feedback (health centres)  - training session, quizzes, and peer-to-peer discussions on the choice of antimicrobial agents using a bag of locally available antibiotics  - introducing treatment guidance for 4 main infections | **1^st^ round:**  Bacterial AMR and its determinants (bacteria and viruses and role of the immune system in combating infection, mechanism how inappropriate antibiotic use induces AMR); Antimicrobial agents, their spectrum of activity, the role of broad vs. narrow spectrum antibiotics, route of administration, dosing, side effects, contra-indications; Indications of antibiotic use, the distinction between Access and Watch antibiotics, and why they are used for different infections; Introduce treatment guidance for cough including bronchitis, community-acquired pneumonia, and diarrhoea or other gastro-intestinal complaints  **2^nd^ round:**  Introduce treatment guidance for acute fever without other discriminating symptoms, malaria, and skin or soft tissue infections. Recap previous guidance.  **3^rd^ round:**  Recap treatment guidance. Provider-patient communication strategies. |
| **Health education campaign (persuasive/ enabling) in communities** | |
| - Mass health education sessions (including video projection, quizzes, theatre plays, commitment ceremonies, distribution of soap)  - Door-to-door visits  - Scheduled meeting with target group  - Ad hoc Q&A with residents  - Role plays at schools, followed by group discussion | **Across rounds:** - Handwashing: the importance of using soap and important hand washing moments - Role of antibiotics to treat infections and effect on AMR - Correct use of antibiotics (e.g. role of compliance, not for any illness, …).  - Role of health centre consultation versus self-medication when ill. - Safe sanitation facilities and practices, including wastewater management. - Safe drinking water sources and safe storage.  **1^st^ round:**  - video projection of existing songs on handwashing - theatre play depicting a dialogue between neighbours on the consequences of drinking water from dams, wells, and of taking medicines - educational group discussions using photovoice images (pictures taken by community members of day-to-day situations driving AMR) - soap distribution and setting up handwashing facilities  **2^nd^ round:** - organise community clean-up days, installation of hand-washing stations - educational campaign on safe sanitation and the consequences of poor adherence to antibiotic treatment - school game on daily routines, highlighting moments of handwashing, install handwashing points in schools - video screening on all topics  **3^rd^ round:**  - repeat all discussed drivers of AMR through all channels; Q&A; commitments on seeking (formal) healthcare consultations when ill |

**Table S 1: Components and content of the co-created community-level AMR intervention bundle.**

| **Domain** | **Indicator (survey question)** | **Category** | **Definition / Response options** |
| --- | --- | --- | --- |
| **Drinking water source** | Access to safe drinking water (Q2 & Q3, binary) | **Improved** | Household tap;  Concession tap;  Public tap;  Borehole;  Finished well;  Rain water;  Water from a bottle |
|  |  | **Unimproved** | Undeveloped well;  Surface water;  Water from a bag |
| **Sanitation** | Using improved sanitary facility (Q7, binary) *Combined with Q9 (shared toilet y/n); shared = unimproved* | **Improved** | Flush toilet connected to a septic tank;  Improved pit latrine with ventilation;  Pit latrine with slab |
|  |  | **Unimproved** | Pit latrine without slab Open defecation |
| **Handwashing** | Correct handwashing – Combination of Q14 (soap available, binary) and Q15 (handwashing before defaecation, binary) | **Correct** | Q14: Yes, bar or liquid soap;  Q15: Yes, always or Yes, often but not always |
|  |  | **Not correct** | Q14: No washing product available;  Washing product available but not used in last 24 h; Ash/mud/sand used;  Detergent (powder/liquid/paste) used;  Q15: No/rarely; Not sure |
| **Animal contact** | Livestock animals access the house (Q17, binary) | **Access** | Yes, inside and outside the house;  Yes, outside but adjoining the house;  Yes, outside but within a defined area |
|  |  | **No access** | No |
| **Animal/ environmental exposure risk** | Animal excrement on floor (Q20, binary) | **Yes** | Yes |
|  |  | **No** | No |

**Table S 2: Binary WASH evaluation framework**

| **All households (n=808)** |  |  |  |  |
| --- | --- | --- | --- | --- |
|  | **Pre-intervention** | | **Post-intervention** | |
| **WASH Indicator** | **Control** | **Intervention** | **Control** | **Intervention** |
|  | (N=409) | (N=399) | (N=393) | (N=383) |
| Main drinking water source (dry) |  |  |  |  |
| Improved | 336 (82.2%) | 382 (95.7%) | 340 (86.5%) | 361 (94.3%) |
| Unimproved | 71 (17.4%) | 16 (4.0%) | 52 (13.2%) | 20 (5.2%) |
| Missing | 2 (0.5%) | 1 (0.3%) | 1 (0.3%) | 2 (0.5%) |
| Main drinking water source (rainy) |  |  |  |  |
| Unimproved | 324 (79.2%) | 367 (92.0%) | 344 (87.5%) | 364 (95.0%) |
| Improved | 83 (20.3%) | 31 (7.8%) | 48 (12.2%) | 17 (4.4%) |
| Missing | 2 (0.5%) | 1 (0.3%) | 1 (0.3%) | 2 (0.5%) |
| Handwashing practices |  |  |  |  |
| Not Correct | 353 (86.3%) | 341 (85.5%) | 326 (83.0%) | 332 (86.7%) |
| Correct** | 54 (13.2%) | 55 (13.8%) | 67 (17.0%) | 48 (12.5%) |
| Missing | 2 (0.5%) | 3 (0.8%) | 0 (0%) | 3 (0.8%) |
| Sanitation facility used |  |  |  |  |
| Unimproved | 266 (65.0%) | 313 (78.4%) | 230 (58.5%) | 292 (76.2%) |
| Improved | 139 (34.0%) | 83 (20.8%) | 160 (40.7%) | 90 (23.5%) |
| Missing | 4 (1.0%) | 3 (0.8%) | 3 (0.8%) | 1 (0.3%) |
| Livestock animals access to the house |  |  |  |  |
| Access | 25 (6.1%) | 49 (12.3%) | 77 (19.6%) | 78 (20.4%) |
| No Access | 382 (93.4%) | 349 (87.5%) | 315 (80.2%) | 304 (79.4%) |
| Missing | 2 (0.5%) | 1 (0.3%) | 1 (0.3%) | 1 (0.3%) |
| Animal excrement on the household floor |  |  |  |  |
| Exposed | 124 (30.3%) | 121 (30.3%) | 175 (44.5%) | 149 (38.9%) |
| Not Exposed | 282 (68.9%) | 273 (68.4%) | 212 (53.9%) | 230 (60.1%) |
| Missing | 3 (0.7%) | 5 (1.3%) | 6 (1.5%) | 4 (1.0%) |
| **Households where stool samples were collected (n = 265)** | | | | |
|  | **Pre-intervention** | | **Post-intervention** | |
| **WASH Indicator** | **Control** | **Intervention** | **Control** | **Intervention** |
|  | (N=134) | (N=132) | (N=131) | (N=127) |
| Main drinking water source (dry) |  |  |  |  |
| Improved | 111 (82.8%) | 127 (96.2%) | 113 (86.3%) | 117 (92.1%) |
| Unimproved | 22 (16.4%) | 5 (3.8%) | 18 (13.7%) | 9 (7.1%) |
| Missing | 1 (0.7%) | 0 (0%) | 0 (0%) | 1 (0.8%) |
| Main drinking water source (rainy) |  |  |  |  |
| Unimproved | 108 (80.6%) | 121 (91.7%) | 115 (87.8%) | 120 (94.5%) |
| Improved | 25 (18.7%) | 11 (8.3%) | 15 (11.5%) | 6 (4.7%) |
| Missing | 1 (0.7%) | 0 (0%) | 1 (0.8%) | 1 (0.8%) |
| Handwashing practices |  |  |  |  |
| Not Correct | 116 (86.6%) | 111 (84.1%) | 109 (83.2%) | 110 (86.6%) |
| Correct** | 17 (12.7%) | 21 (15.9%) | 22 (16.8%) | 16 (12.6%) |
| Missing | 1 (0.7%) | 0 (0%) | 0 (0%) | 1 (0.8%) |
| Sanitation facility used |  |  |  |  |
| Unimproved | 89 (66.4%) | 102 (77.3%) | 75 (57.3%) | 91 (71.7%) |
| Improved | 44 (32.8%) | 30 (22.7%) | 54 (41.2%) | 35 (27.6%) |
| Missing | 1 (0.7%) | 0 (0%) | 2 (1.5%) | 1 (0.8%) |
| Livestock animals access to the house |  |  |  |  |
| Access | 13 (9.7%) | 19 (14.4%) | 33 (25.2%) | 30 (23.6%) |
| No Access | 120 (89.6%) | 113 (85.6%) | 98 (74.8%) | 96 (75.6%) |
| Missing | 1 (0.7%) | 0 (0%) | 0 (0%) | 1 (0.8%) |
| Animal excrement on the household floor |  |  |  |  |
| Exposed | 42 (31.3%) | 42 (31.8%) | 64 (48.9%) | 57 (44.9%) |
| Not Exposed | 91 (67.9%) | 89 (67.4%) | 67 (51.1%) | 68 (53.5%) |
| Missing | 1 (0.7%) | 1 (0.8%) | 0 (0%) | 2 (1.6%) |

**Table S 3:** **WASH household survey results.** Percentage of households reporting unfavourable WASH exposures by study group and survey round in the household cohort among those households where a WASH survey only was conducted (A) and among households where a WASH survey and stool samples were collected (B).

| **Round** | **Strata** | **Esbl-E** | **n** | **N** | **Prevalence** |
| --- | --- | --- | --- | --- | --- |
| 1 | Health centre (government run) | 0 | 322 | 693 | 0.46 |
| 1 | Health centre (government run) | 1 | 371 | 693 | 0.54 |
| 1 | No health centre (other informal/formal) | 0 | 244 | 510 | 0.48 |
| 1 | No health centre (other informal/formal) | 1 | 266 | 510 | 0.52 |

**Table S 4. Baseline ESBL-E prevalence by randomization stratum.** Stratified by presence of a government-run health centre. n/N indicates number positive over total sampled (prevalence)**.**

| **Scenario** | **Parameter** | **Median (95%CrI)** |
| --- | --- | --- |
| S1 | between-household (SD) | 1.349 (1.065 - 1.668) |
| S1 | between-household (variance) | 1.820 (1.134 – 2.783) |
| S2 | between-household (SD) | 1.353 (1.072 – 1.672) |
| S2 | between-household (variance) | 1.830 (1.149 – 2.796) |

**Table S 5: Posterior estimates of between-household variability in transition hazards.** Posterior median and 95% credible intervals (CrI) for the standard deviation and variance of the household-level random effects on the log-hazard scale, estimated from the Bayesian hierarchical model under each scenario.

| **Village** | **Group** | **ESBL–E acquisition rate Baseline  Median (95%CrI)  per 100 person–days** | **IRR Early phase** | **IRR  Late phase** |
| --- | --- | --- | --- | --- |
| 1 | Intervention | 0.14 (0.06–0.89) | 1.02 (0.77–1.36) | 0.79 (0.51–1.19) |
| 2 | Intervention | 4.01 (1.61–15.39) | 0.95 (0.69–1.29) | 0.76 (0.50–1.14) |
| 3 | Control | 3.14 (1.22–13.45) | - | - |
| 4 | Intervention | 2.84 (1.08–11.51) | 0.99 (0.72–1.37) | 0.79 (0.49–1.25) |
| 5 | Control | 3.07 (1.24–12.29) | - | - |
| 6 | Control | 2.99 (1.22–11.33) | - | - |
| 7 | Intervention | 3.53 (1.47–14.34) | 0.88 (0.55–1.61) | 0.70 (0.39–1.41) |
| 8 | Intervention | 3.17 (1.31–12.02) | 0.99 (0.72–1.36) | 0.80 (0.51–1.20) |
| 9 | Control | 3.28 (1.32–12.61) | - | - |
| 10 | Control | 3.62 (1.47–13.98) | - | - |
| 11 | Intervention | 3.53 (1.4–14.35) | 1.01 (0.76–1.36) | 0.80 (0.46–1.30) |
| 12 | Intervention | 3.71 (1.49–13.68) | 0.91 (0.58–1.33) | 0.71 (0.37–1.15) |
| 13 | Intervention | 2.94 (1.19–11.92) | 1.01 (0.77–1.32) | 0.77 (0.49–1.17) |
| 14 | Control | 4.66 (1.92–17.08) | - | - |
| 15 | Intervention | 2.57 (0.95–17.81) | 0.90 (0.59–1.56) | 0.74 (0.43–1.37) |
| 16 | Control | 3.25 (1.34–12.45) | - | - |
| 17 | Control | 2.63 (1.01–11.03) | - | - |
| 18 | Control | 3.29 (1.34–12.6) | - | - |
| 19 | Control | 3.22 (1.27–14.26) | - | - |
| 20 | Control | 4.26 (1.76–16.1) | - | - |
| 21 | Intervention | 3.61 (1.47–13.95) | 0.96 (0.70–1.29) | 0.74 (0.47–1.15) |
| 22 | Intervention | 3.06 (1.25–11.73) | 1.00 (0.68–1.50) | 0.81 (0.50–1.32) |

**Table S 6:** **Incidence rate ratio per intervention village cluster compared to calendar-matched controls.** The IRR are computed against calendar-matched pooled controls. Calendar-matched control = pooled across all calendar-match control villages, calendar-time matched to each intervention village and intervention phase. IRRs rather than HRs are presented at village-level as the model did not directly estimate village-specific hazards (i.e. we incorporated clustering at household-, not village-level). We estimated village-level incidences during the different phases, by predicting the incidence based on the population characteristics of each village (age, sex, and household-level effects) and compared that to a pooled control group.

| **Scenario** | **Metric** | **P(reduction) (HR/IRR < 1)** | **P(increase) (HR/IRR > 1)** | **P(meaningful reduction) (HR/IRR < 0.9)** | **P(trivial reduction) (0.9 ≤ HR/IRR ≤ 1)** | **P(meaningful increase) (HR/IRR > 1)** | **P(within ROPE 0.9–1.1)** |
| --- | --- | --- | --- | --- | --- | --- | --- |
| S1 | HR_phase1 | 45 | 55 | 18 | 27 | 55 | 54.3 |
| S1 | HR_phase2 | 88 | 12 | 71.1 | 16.9 | 12 | 25.1 |
| S1 | IRR_phase1 (DiD) | 62.3 | 37.8 | 38.1 | 24.2 | 37.8 | 42.1 |
| S1 | IRR_phase2 (DiD) | 84.1 | 15.9 | 71.4 | 12.8 | 15.9 | 20.5 |
| S1 | IRR_overall (DiD) | 70.8 | 29.2 | 45.8 | 25 | 29.2 | 40.5 |
| S2 | HR_phase1 | 74 | 26 | 36.6 | 37.3 | 26 | 57.5 |
| S2 | HR_phase2 | 74 | 26 | 36.6 | 37.3 | 26 | 57.5 |
| S2 | IRR_phase1 (DiD) | 81.7 | 18.3 | 58.9 | 22.8 | 18.3 | 34.1 |
| S2 | IRR_phase2 (DiD) | 71.1 | 28.9 | 49.9 | 21.3 | 28.9 | 35.3 |
| S2 | IRR_overall (DiD) | 79.9 | 20.1 | 56.7 | 23.3 | 20.1 | 35.2 |

**Table S 7: Posterior probabilities for intervention effects on ESBL-E transmission dynamics.** Values show the percentage of posterior draws indicating a reduction (HR/IRR < 1), increase (HR/IRR > 1), meaningful reduction (HR/IRR < 0.9), trivial reduction (0.9–1), and within the ROPE (0.9–1.1; negligible effects). HRs represent acquisition hazards and IRRs represent cumulative incidence. S1: time-varying effects; S2: constant effect.

| **Model scenario** | **Intervention  scenarios** | **Seasonal effect** | **Intervention effect** | **LooIC** | **LooIC** | **Rhat** | **Rhat** | **N.div.** |
| --- | --- | --- | --- | --- | --- | --- | --- | --- |
|  |  |  |  |  | **diff** | **min** | **max** |  |
| 1A (Base case) | Two step | Sine | Acquisition | 3783 | 2.8 | 1 | 1.003 | 0 |
| 2A | One step | Sine | Acquisition | 3784 | 3 | 1 | 1.003 | 1 |
| 1B | Two step | Spline | Acquisition | 3785 | 4 | 1 | 1.011 | 3 |
| 2B | One step | Spline | Acquisition | 3786 | 5.8 | 1 | 1.017 | 40 |
| 3A | Two step | Sine | Acquisition + decolonisation | 3790 | 9 | 1 | 1.002 | 0 |
| 3B | Two step | Spline | Acquisition + decolonisation | 3784 | 3 | 1 | 1.007 | 234 |
| 4B | One step | Spline | Acquisition + decolonisation | 3781 | 0 | 1 | 1.011 | 343 |
| 4A | One step | Sine | Acquisition + decolonisation | 3794 | 2 | 1 | 1.002 | 2 |
| 1C | Two step | None | Acquisition | 3841 | 60.4 | 1 | 1.002 | 13 |

**Table S 8:** **Comparative model fits.** LooIC = leave-on-out information criterion. The smaller the value, the better the fit. N div = number of divergences. Divergences result in less reliable results. Base case = baseline scenario.

**CONSORT 2010 checklist of information to include when reporting a cluster randomised trial**

| **Section/Topic** | Item No | Standard Checklist item | Extension for cluster designs | Page No * |
| --- | --- | --- | --- | --- |
| **Title and abstract** | | | |  |
|  | 1a | Identification as a randomised trial in the title | Identification as a cluster randomised trial in the title | 1 |
|  | 1b | Structured summary of trial design, methods, results, and conclusions (for specific guidance see CONSORT for abstracts) | See table 2 | 2 |
| **Introduction** | | | |  |
| **Background and objectives** | 2a | Scientific background and explanation of rationale | Rationale for using a cluster design | 5-6 |
|  | 2b | Specific objectives or hypotheses | Whether objectives pertain to the cluster level, the individual participant level or both | 5-6 |
| **Methods** | | | |  |
| **Trial design** | 3a | Description of trial design (such as parallel, factorial) including allocation ratio | Definition of cluster and description of how the design features apply to the clusters | 6 |
|  | 3b | Important changes to methods after trial commencement (such as eligibility criteria), with reasons |  | NA |
| **Participants** | 4a | Eligibility criteria for participants | Eligibility criteria for clusters | 7 |
|  | 4b | Settings and locations where the data were collected |  | 6-8 |
| **Interventions** | 5 | The interventions for each group with sufficient details to allow replication, including how and when they were actually administered | Whether interventions pertain to the cluster level, the individual participant level or both | 6-7 and  Table S1 |
| **Outcomes** | 6a | Completely defined pre-specified primary and secondary outcome measures, including how and when they were assessed | Whether outcome measures pertain to the cluster level, the individual participant level or both | 8 |
|  | 6b | Any changes to trial outcomes after the trial commenced, with reasons |  | NA |
| **Sample size** | 7a | How sample size was determined | Method of calculation, number of clusters(s) (and whether equal or unequal cluster sizes are assumed), cluster size, a coefficient of intracluster correlation (ICC or *k*), and an indication of its uncertainty | 9 |
|  | 7b | When applicable, explanation of any interim analyses and stopping guidelines |  | NA |
| **Randomisation:** | | | |  |
| **Sequence generation** | 8a | Method used to generate the random allocation sequence |  | 6 and Joint submission |
|  | 8b | Type of randomisation; details of any restriction (such as blocking and block size) | Details of stratification or matching if used | 6 |
| **Allocation concealment mechanism** | 9 | Mechanism used to implement the random allocation sequence (such as sequentially numbered containers), describing any steps taken to conceal the sequence until interventions were assigned | Specification that allocation was based on clusters rather than individuals and whether allocation concealment (if any) was at the cluster level, the individual participant level or both | 6 and joint submission |
| **Implementation** | 10 | Who generated the random allocation sequence, who enrolled participants, and who assigned participants to interventions | Replace by 10a, 10b and 10c | 6 and joint submission |
|  | 10a |  | Who generated the random allocation sequence, who enrolled clusters, and who assigned clusters to interventions | 6 and joint submission |
|  | 10b |  | Mechanism by which individual participants were included in clusters for the purposes of the trial (such as complete enumeration, random sampling) | 6 and joint submission |
|  | 10c |  | From whom consent was sought (representatives of the cluster, or individual cluster members, or both), and whether consent was sought before or after randomisation | 7-8 |
| **Blinding** | 11a | If done, who was blinded after assignment to interventions (for example, participants, care providers, those assessing outcomes) and how |  | NA |
|  | 11b | If relevant, description of the similarity of interventions |  | NA |
| **Statistical methods** | 12a | Statistical methods used to compare groups for primary and secondary outcomes | How clustering was taken into account | 9-10 |
|  | 12b | Methods for additional analyses, such as subgroup analyses and adjusted analyses |  | 10 |
| **Results** | | | |  |
| **Participant flow (a diagram is strongly recommended)** | 13a | For each group, the numbers of participants who were randomly assigned, received intended treatment, and were analysed for the primary outcome | For each group, the numbers of clusters that were randomly assigned, received intended treatment, and were analysed for the primary outcome | Joint submission and Figure 1 |
|  | 13b | For each group, losses and exclusions after randomisation, together with reasons | For each group, losses and exclusions for both clusters and individual cluster members | Joint submission and Figure 1 |
| **Recruitment** | 14a | Dates defining the periods of recruitment and follow-up |  | 7-8 |
|  | 14b | Why the trial ended or was stopped |  | NA |
| **Baseline data** | 15 | A table showing baseline demographic and clinical characteristics for each group | Baseline characteristics for the individual and cluster levels as applicable for each group | Table 1  Table S4 |
| **Numbers analysed** | 16 | For each group, number of participants (denominator) included in each analysis and whether the analysis was by original assigned groups | For each group, number of clusters included in each analysis | Figure 1 |
| **Outcomes and estimation** | 17a | For each primary and secondary outcome, results for each group, and the estimated effect size and its precision (such as 95% confidence interval) | Results at the individual or cluster level as applicable and a coefficient of intracluster correlation (ICC or k) for each primary outcome |  |
|  | 17b | For binary outcomes, presentation of both absolute and relative effect sizes is recommended |  | 13-14 |
| **Ancillary analyses** | 18 | Results of any other analyses performed, including subgroup analyses and adjusted analyses, distinguishing pre-specified from exploratory |  | 14 |
| **Harms** | 19 | All important harms or unintended effects in each group (for specific guidance see CONSORT for harms) |  | NA/ Joint submission |
| **Discussion** | | | |  |
| **Limitations** | 20 | Trial limitations, addressing sources of potential bias, imprecision, and, if relevant, multiplicity of analyses |  | 17 |
| **Generalisability** | 21 | Generalisability (external validity, applicability) of the trial findings | Generalisability to clusters and/or individual participants (as relevant) | 15-17 |
| **Interpretation** | 22 | Interpretation consistent with results, balancing benefits and harms, and considering other relevant evidence |  | 15-17  4 |
| **Other information** | | |  |  |
| **Registration** | 23 | Registration number and name of trial registry |  |  |
| **Protocol** | 24 | Where the full trial protocol can be accessed, if available |  | 18 |
| **Funding** | 25 | Sources of funding and other support (such as supply of drugs), role of funders |  | 18 |

**Table S 8. CONSORT checklist.** * Note: page numbers optional depending on journal requirements
